# Copy-number alterations reshape the classification of diffuse intrinsic pontine gliomas. First exome sequencing results of the BIOMEDE trial

**DOI:** 10.1101/2021.04.29.21256183

**Authors:** Thomas Kergrohen, David Castel, Gwenael Le Teuff, Arnault Tauziède-Espariat, Emmanuèle Lechapt-Zalcman, Karsten Nysom, Klas Blomgren, Pierre Leblond, Anne-Isabelle Bertozzi, Emilie De Carli, Cécile Faure-Conter, Celine Chappe, Natacha Entz-Werlé, Angokai Moussa, Samia Ghermaoui, Emilie Barret, Stephanie Picot, Marjorie Sabourin-Cousin, Kevin Beccaria, Gilles Vassal, Pascale Varlet, Stephanie Puget, Jacques Grill, Marie-Anne Debily

## Abstract

Diffuse intrinsic pontine gliomas (DIPG) is an incurable neoplasm occurring mainly in children for which no progress was made in the last decades. The randomized phase II BIOMEDE trial compared three drugs (everolimus, dasatinib, erlotinib) combined with irradiation. The present report describes whole exome sequencing (WES) results for the first 100 patients randomized.

Copy-number-Alteration (CNA) unsupervised clustering identified four groups with different outcomes and biology. This classification improved prognostication compared to models based on known biomarkers (Histone H3 and *TP53* mutations). The cluster presenting complex genomic rearrangements was associated with significantly worse outcome and TP53 dysfunction. Mutation and CNA signatures confirmed the frequent alteration in DNA repair machinery. With respect to potential targetable pathways, PI3K/AKT/mTOR activation occurred in all the samples through multiples mechanisms. In conclusion, WES at diagnosis was feasible in most patients and provides a better patient stratification and theranostic information for precision medicine.

## Introduction

Pediatric high-grade gliomas (HGG) represent the most frequent malignant brain tumors in children and one of the main causes of cancer-related death in this age group. These neoplasms are biologically different from those encountered in the adult population (1) and this may explain the lack of efficacy of the standard of care established for adult with high-grade glioma (2,3). Diffuse Intrinsic Pontine Glioma (DIPG), especially, exhibit in most cases a unique mutation (K27M) in the regulatory tail of H3 histone impairing PRC2 function, hereby modifying deeply the epigenetic landscape of the tumor cells (4,5). This mutation is specific to most Diffuse Midline Gliomas in children, adolescent and young adults and defines now a new entity recognized by the 2016 update of the WHO classification (6). DIPG represent the archetypal form of these neoplasms and their treatment has been individualized for decades in specific protocols (7). Radiotherapy is the only treatment with proven efficacy but recurrence occurs inevitably after a few months; none of the medical treatment tested so far has been able to extend survival (7–9).

In recent years, autopsies and biopsies at diagnosis could describe more precisely the molecular landscape of DIPG (1,10–13). Four subtypes of DIPG were identified based on the mutations in gene encoding the canonical histone protein H3.1/H3.2 or variant H3.3 (14) and more recently on the overexpression of *EZHIP* or *EGFR* mutation in rare H3-wildtype tumors (15,16). Also, several potential therapies have been identified in preclinical models, some of them being introduced in precision medicine trials (17,18). The BIOMEDE (Biological Medicine for DIPG Eradication) trial (NCT02233049) was designed to explore the feasibility of biology-driven targeted therapy combined with irradiation in this setting. After histological confirmation of the diagnosis with a stereotactic biopsy, patients were allocated to one of three treatment arms (erlotinib, everolimus or dasatinib) on the basis of the presence of their known target(s) in the tumor. In the case where more than one target was present, patients were randomized. Tumor profiling with whole-exome sequencing (WES) was performed with the aim to identify additional actionable therapeutic alterations that could be targeted at relapse. We present here the results of the first 100 tumors that were profiled in the study.

## Results

### Patient population

In the first three and a half years of the trial (October 6th, 2014 – January 19th, 2018), one hundred and twenty-four patients were enrolled and had a biopsy at diagnosis. Central pathology review confirmed the diagnosis in 116 patients presenting a global loss of H3K27me3 epigenetic mark along with H3K27M mutation or *EZHIP* overexpression. Eight patients (7%) were excluded as diagnosis was reclassified to: *MYCN* high-grade gliomas (2), ganglioglioma with H3K27M and BRAF-V600E mutation (1), angiocentric glioma with *QKI* translocation (1), NF1-associated glioma (1), pilomyxoïd astrocytoma (1), and glioma, NOS with too limited material to confirm the DIPG diagnosis made locally (2). None did correspond to Diffuse Midline Glioma (DMG) with *EGFR* alteration recently described(16).

### Most DIPG stereotactic biopsies yielded high quality WES data

Out of the 116 patients with a confirmed DIPG diagnosis, frozen material was available for 104 cases, but matched blood sample was missing and DNA yields were too low for 6 and 3 cases, respectively (Supplementary Fig. S1A). Ninety-five tumors could therefore be analysed by WES (95/104 cases *i*.*e*. 91%), and an average of 170 and 122 million reads for tumor and blood samples were collected, respectively. The tumor cell content could be estimated based on the variant allele frequency (VAF) of the heterozygous H3K27M driver mutation in patients harboring *H3F3A* or *HIST1H3B* alterations in the absence of copy number changes at the locus of the histone mutated. The site of the biopsy was located for 39 patients according to the hypersignal on T2/FLAIR on postoperative MR imaging as: inside the tumor in 17, at the edge in 14 and outside in 8 (Fig. 1A-C). Despite a slight trend, there was no significant correlation between the site of the biopsy and the estimated tumor cell content (Kruskal-Wallis, p=0.42, Fig. 1D). Also, the tumor cell content did not influence the mutational burden identified with different variant callers (Fig. 1E & Supplementary Fig. S1C, p=0.07 & 0.8) reflecting that a lower percentage of tumor cells did not impair variant detection.

**Figure 1:**
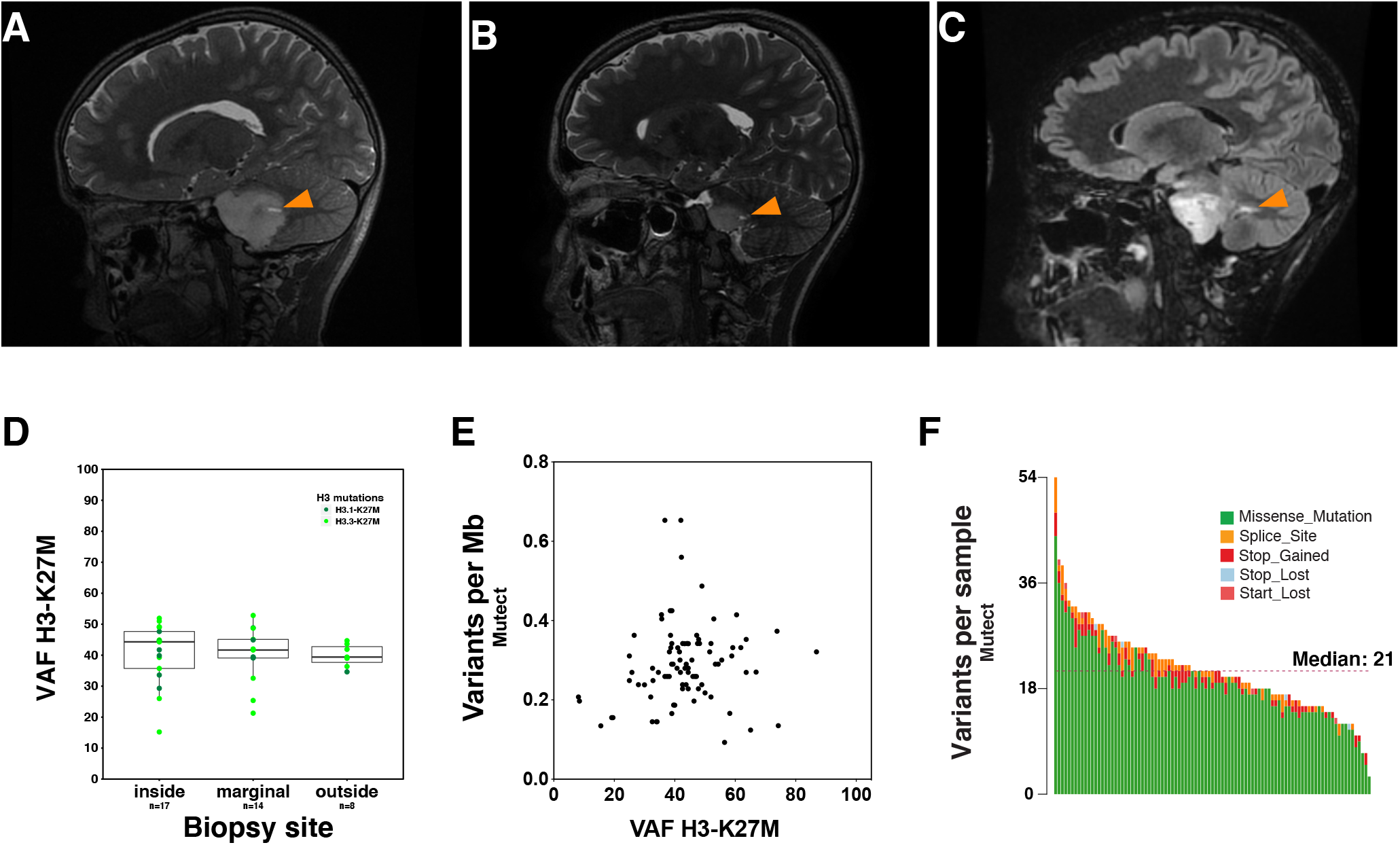
Impact of biopsy location on tumor burden. Representative MRI images for biopsies with a location inside (A), on the edge (B) and outside (C) the tumor as shown by orange arrows. Influence of biopsy site on sample tumor content (D). The Variant Allele Frequency of the H3K27M mutation is on average higher for ‘inside tumor’ biopsies, but the difference in distribution is not significative (Kruskal-Wallis, *p=* 0.42). Relationship between sample tumor content and mutation load from Mutect variant caller analysis (E). The number of single nucleotide variations per Mb detected using Mutect is reported according to the Variant Allele Frequency of the H3K27M mutation as an approximation of the tumor content in patient harboring either *H3F3A* or *HIST1H3B* mutations. No significant correlation was identified (Spearman correlation 0.1956, p=0.069). Stacked barplot representing the number and types of variants in each sample detected by Mutect variant calling (F).

The mutation rate fluctuated importantly among patients independently of the H3 mutational status, but the number and distribution of the type of non-synonymous coding variants was mostly similar to previous reports using variant caller as stringent as Mutect with a median of 21 mutations per exome for each patient (ranging from 3 to 51) (1,19)(Fig. 1F & Supplementary Fig. S1B-D). We also used Varscan caller to detect small indel variations and to include them in subsequent recurrence analysis increasing the median of variations to 24 per patient (ranging from 8 to 61).

### DIPG somatic CNA landscape distinguishes 4 different clusters

We analyzed genome-wide copy number alterations (CNAs) and identified by unsupervised hierarchical clustering four clusters of DIPG harboring distinct profiles (Fig.2, Supplementary Fig. S2A-B). Samples from cluster 1 (C1) and 2 (C2) were associated with high CNA levels defined as the percentage of the genome bearing copy number gains or losses. All C1 samples presented complex genomic rearrangements distributed throughout the genome with on average 32% of the genome with chromosomal imbalance (Supplementary Fig. S2B). C2 samples are characterized by chromosome 1q and 2 gains, with a complex CNA profile similar to C1 samples for around one third of cases. The others genomic profiles were associated with a low CNA burden and almost no CNA in cluster 3 (C3), and an overall good genome integrity except chromosome 1q gain in cluster 4 (C4) (Fig.2, Supplementary Fig. S2A-B). The detection of fewer CNA in samples from C3 did not solely result from a weak number of tumor cells in the biopsy as the number of mutations per Mb was at least similar to samples presenting chromosome 1q gains in C4.

**Figure 2:**
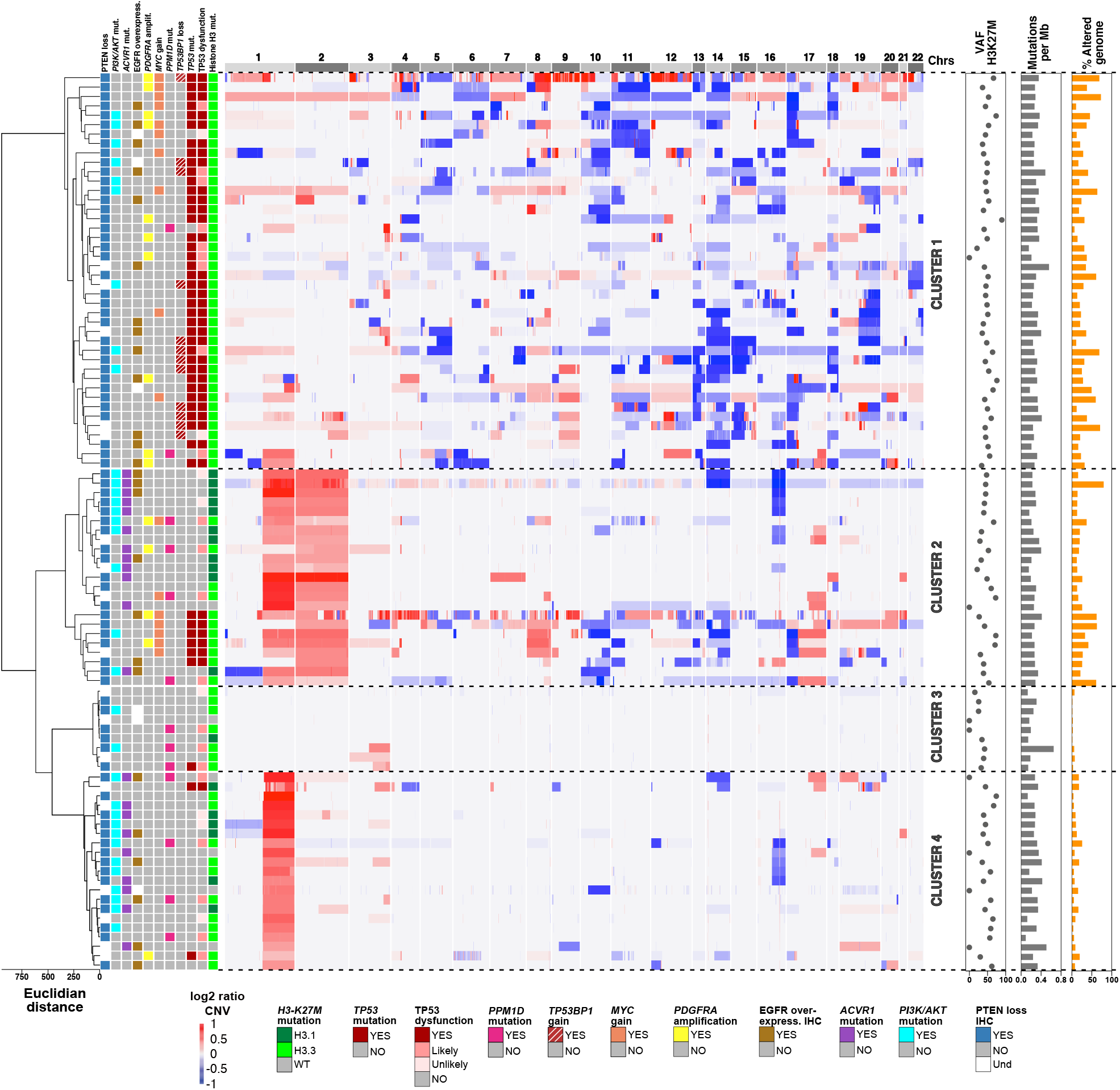
Hierarchical clustering of Copy Number Alterations in DIPG. Unsupervised hierarchical clustering and heatmap representing the copy number genomic alterations of the 95 DIPG patients profiled by WES (red, gain; blue, loss reflecting the log2 ratio of tumor *vs*. blood copy number values). Samples are arranged in rows and chromosomes in columns. Clinicopathological (PTEN loss in immunohistochemistry) and molecular annotations are provided as bars on the left side according to the color code indicated below the heatmap. Coding SNVs associated with a VAF = or > to 5%, a minimum of 5 supporting reads and a frequency below 1% in 1000G DB are reported as well as *53BP1* loss. The VAF of H3K27M mutation is indicated as a scatter dot plot as well as the tumor mutation load (*i*.*e*. total number of SNV) from Mutect analysis and the proportion of the genome presenting CNA variations as histograms on the right.

We then aimed to characterize each of these clusters. The proportion of patients below the age of 5 years at diagnosis was significantly higher in clusters presenting few genomic alterations, *i*.*e*. C3 and C4 (>44%, p=0.022, Fig. 3). Distribution of H3 mutational status was not random as C1 contained only H3.3 mutated samples (p<0.0001), yet the clustering did not reflect purely the histone H3 gene mutated (Fig. 2-3). Indeed, H3.1-K27M samples were scattered in C2, 3 & 4, containing also many H3.3 mutated and H3 wild-type tumors.

**Figure 3:**
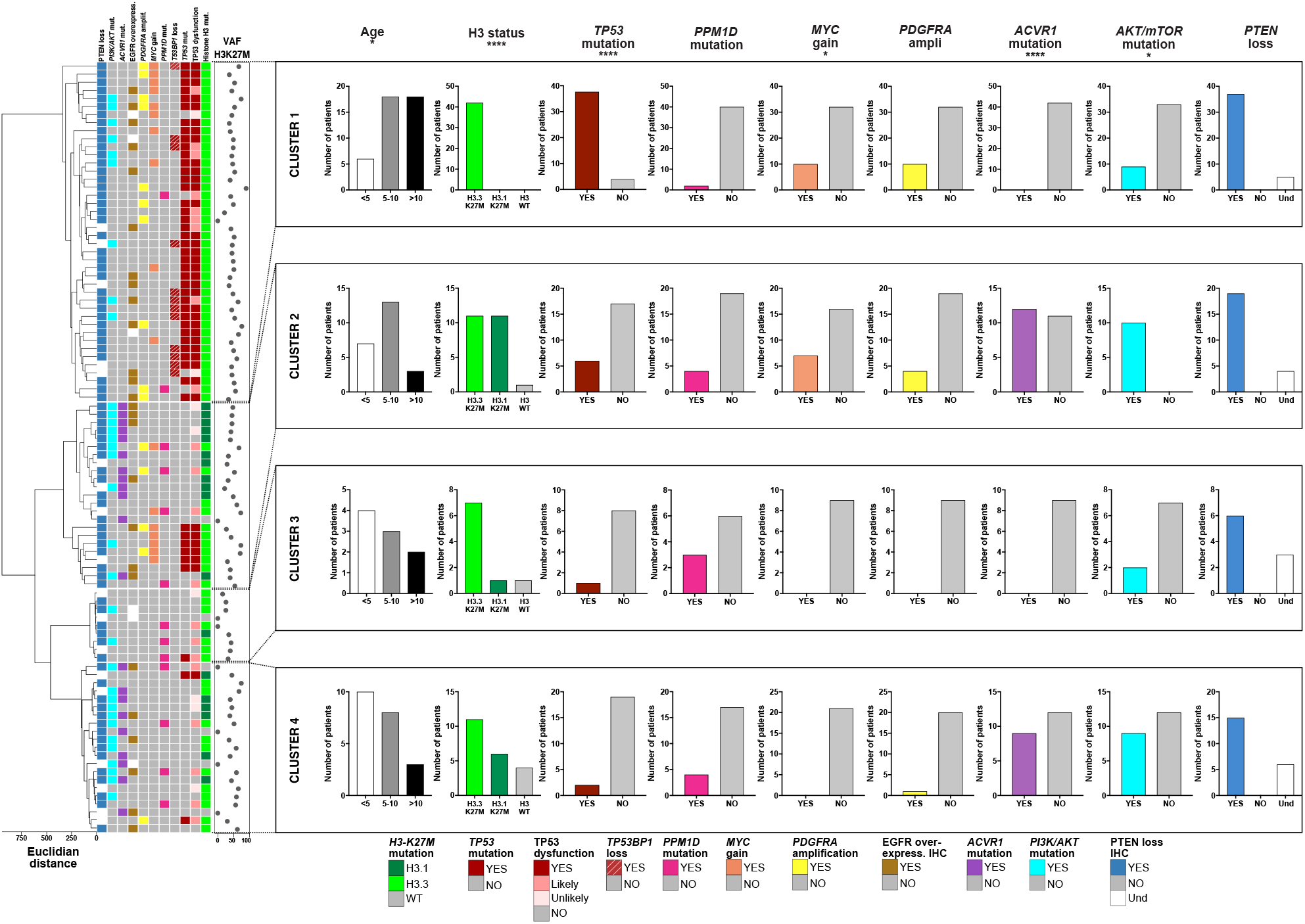
Molecular and clinicopathological description of DIPG CNA subgroups. Dendrogram resulting from the unsupervised hierarchical clustering of CNA shown in fig2 is presented concomitantly with the distribution of clinicopathological and molecular information within the 4 clusters. Fisher test, \*\*\*\**p* <0.0001, ** *p* <0.01. *p*-value: age 0.024, H3 status 4e-6, *TP53* mutation 3.0e-14, *PPM1D* mutation 0.28, *MYC* gain 3.4e-10, *PDGFRA* amplification 0.003, *ACVR1* mutation 5.8e-07, AKT/mTOR mutation 0.0039, *PTEN* loss 0.2756).

Among specific recurrent alterations, a significant difference of distribution was observed for *TP53* (p=3.05e-11) enriched in cluster 1 and 2, *ACVR1* (p=5.762e-07) restricted to C2 and 4, *AKT/MTOR* pathway mutations enriched in C2 and 4 (p=0.003943) and *PDGFRA* amplification enriched in C1 & 2 (p=0.003514) (Fig. 3 and Supplementary Fig. S2C). PTEN loss was present in all the tumors analysed by immunohistochemistry with positive internal control on the vessels (n=77). In a subset of samples, pS6 and pAKT were also found positive in most of the cases (*data not shown*). Overexpression of EGFR was detected in 29% of patients from all clusters but C3 (p=0.0621), maybe resulting from the smaller size of this subgroup. We did not identify a molecular characteristic associated with C3 samples lacking CNA.

Analysis of cluster-specific recurrent chromosomal imbalance of genomic regions retrieved the genes found mutated or associated with CNA in H3.3-K27M samples by Mackay *and coll*.*(1), i*.*e*. gains of *PDGFRA* in C1&2, *CCND1, CCND2, IGF1R* and *TOP3A* in C1, *KIT* and *KDR* in C2, losses of *AURKB, KDM6B* and *TP53* and in C1. However, gains of *CDK6* (ampli n=1, gain n=4), *EGFR* (gain n=4) and *MET* (ampli n=2, gain n=4) found in restricted numbers of patients in our cohort were not selected by GISTIC as *MYC* amplification observed in only two patients and moderate gains in 15 additional ones (Supplementary Fig. S2C). We did not retrieve either any recurrent *CDKN2A/CDKN2B* deletion. In addition, we highlighted frequent loss of *TP53BP1* (n= 12, 28%) specific to C1 samples and *SUFU* (n=8, 35%), *TLX1* and *TRIM8* in cluster C2. *SUFU* is regulating the Sonic Hedgehog pathway which has been implicated in the oncogenesis of DIPG in earlier studies (20). *TLX1* is involved in early differentiation of the mammalian CNS. *TRIM8* could regulate stemness of GBM GSC and frequent deletion or LOH were reported in adult GBM (21,22).

C1 cluster was the most distant subgroup of samples from all others as highlighted by the dendrogram (Fig.2), and the most homogeneous containing almost exclusively H3.3-K27M, *TP53*-mutated samples (38/42; Fig2-3). Among the four patients without *TP53* mutations, two harbored a truncating mutation in exon 6 of *PPM1D* known to mimic TP53 deficiency in the DNA damage response pathway and could consequently be associated with a ‘*likely’* TP53 dysfunction (23). The last two patients were associated with a loss of heterozygosity in *TP53* and tagged *‘unlikely TP53 dysfunction’* (Fig. 2). However, one of these two DIPG patients also harbored a copy number loss of *TP53BP1* encoding a checkpoint protein which serves as a DNA double strand break sensor whose deletion may play an important role in chromosomal instability as this protein is important for NHEJ DNA repair (24,25). The second patient presented an additional stop-gain mutation (VAF 46.72%) in *NELFCD* gene, a TP53-transcriptional target of the NELF complex regulating BRCA1 recruitment to DNA breakage sites for DNA repair (26).

### Copy number alterations signatures reflect chromosomal instability in DIPG

We then looked for signatures derived from copy number features and estimated, using the cophenetic distances, that the optimal number of CNA signatures was 3 (Supplementary Fig. S3A). The relative proportion of each signature varied from one cluster to the other. DIPG genomes were shaped by at least two dysregulated processes (Supplementary Fig. S3B). The copy number signature 2 reflected a stable genome and accounted for the major contribution in patients from cluster 3; in comparison, the signatures 1 and 3 presented a higher number of breakpoints by arm as well as smaller length of DNA segments, thus reflecting altered genomes (Supplementary Fig. S3A). CN signature 1 displayed a high copy number state and a significant proportion of CN change points of 2 or more, reflecting polyploidy. We noted an enrichment of two breakpoints per chromosome arm that could suggest that the mutational process underlying this signature might correspond to breakage–fusion–bridge (BFB) events (27,28). A significant proportion of CN change point (CNCP) of size one and a CN switch from diploid to single copies, both characteristic of chromothripsis events, were associated with signature 3 Supplementary Fig. S3B). Indeed, the CN of the involved chromosome arms usually oscillates between the normal and deleted CN states and frequent LOH are observed in the low-DNA copy number regions. Accordingly, a significantly higher length of oscillating CN segment chains was observed in signature 3.

We then correlated CNA signatures to molecular features (Supplementary Fig. S3C) and showed positive association of signature 3 with *TP53* mutations, *TP53BP1* loss, chromothripsis score, sample location in cluster 1 and, to a lesser extent, to the mutation load, and a negative association with chromosome 1q gain. Signature 1 showed a positive association with chromosome 2 gain.

### TP53 mutational spectrum in DIPG

Considering the frequent mutation of *TP53* in DIPG and its possible link with CNA, we analysed specifically the distribution, the consequences and the type of alterations encountered in the cohort. *TP53* alterations were present predominantly in cluster 1 (90% of the samples) and to a lower extent in cluster C2 (26%) but only seldomly in C3 and C4 (Fig. 3). Noticeably, the few *TP53*-mutated tumors in C2 were the subset of genomic profiles presenting complex rearrangements (Fig. 2). The percentage of genome altered in DIPG tumors was significantly higher in *TP53*^*MUT*^ tumors in comparison with *TP53*^*WT*^ (median 31% *vs*. 12%, p<0.0001) whereas the mutation load was only slightly higher in *TP53*^*MUT*^ tumors (median 0.29 Mut/Mb in *TP53*^*MUT*^ and 0.27 Mut/Mb in *TP53*^*WT*^, Kolmogorov-Smirnov test p<0.047, Supplementary Fig. S4A).

We then looked at the *TP53* mutational spectrum in DIPG (Supplementary Fig. S4C&D). Concerning the location of somatic *TP53* mutations, they were in all patients but three in the DNA binding domain and therefore likely to exert a dominant negative effect (29). The most frequent mutations corresponded to hotspot missense mutations affecting residue 175, 245, 248, 273 within the DNA binding domain (Supplementary Fig. S4D). Around thirty-six % of patients presented a LOH associated with a point mutation or indel and 8.4% presenting heterozygous compound mutations, all suspected to be associated with TP53 dysfunction, except if the alteration appeared clearly subclonal and were therefore tagged ‘likely TP53 dysfunction’. TP53 dysfunction was supposed to be ‘unlikely’ in tumors with only LOH, frameshift or intronic SNV of *TP53* and ‘likely’ if tumor harbored *PPM1D* alteration (Supplementary Fig. S4C).

*TP53* mutations appeared to be among the earliest events occurring in DIPG because these mutations exhibit some of the highest VAF encountered, irrespective of LOH (median 0.70, 75% CI [0.42-0.84]; Figure 5B).

TP53 alterations appeared to be linked to complex genomic rearrangements as the vast majority of patients with TP53 dysfunction are found in clusters C1 & C2 (Supplementary Fig. S4C) and associated globally with a higher proportion of their genome altered (Supplementary Fig. S4A&C, *p*<0.0001). The patients with an elevated CNA burden (>45% of altered genome) always present a LOH on one *TP53* allele, excepted for one patient without any *TP53* mutation in cluster C2 for which the percentage of genome altered is overestimated because of a noisy baseline on CNA profile. Indeed, for this latter case, detailed examination of CNA profile showed a global good integrity of DNA with only whole chromosome arm gains (chrs 1q & 2) and losses (chrs 14&16).

H3.3-K27M/*TP53* double-mutated patients displayed a significant higher number of segments in particular on chromosomes 8, 14, 17 and 19 suggesting chromothripsis (Fig. 4B).

**Figure 4:**
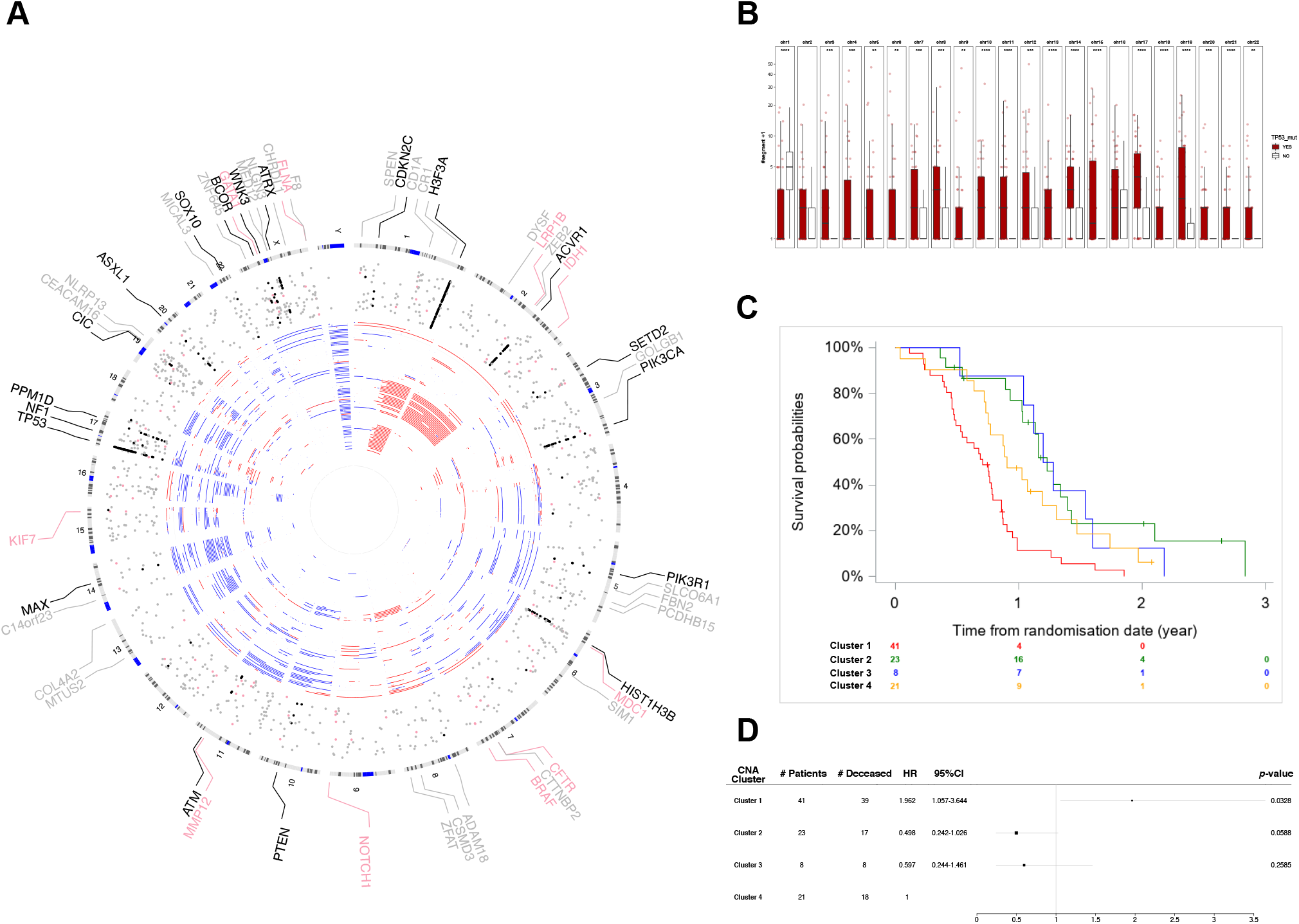
DIPG displaying genome instability are associated with worse prognosis. A-Circos plot depicting the molecular landscape in DIPG. Outer ideogram runs clockwise from chromosome 1 to chromosome Y. Circular tracks from inside to outside: positions of CNA by patient (red: gain, blue: loss); positions of coding somatic SNVs (dots). Because of large number of mutations only genes already described in DIPG with somatic mutation are shown as black dots (Mackay *et al)* or cancer-related genes (known TSG & oncogenes) as pink dots and grey dots otherwise. For this last set of genes, gene name is indicated if mutated in at least 2 distinct patients in our cohort. B – Box plot reflecting the distribution of the number of segments detected by CNV analysis of WES data according to the mutational status of *TP53*. The value number of segments +1 is plotted in log10 scale. Wilcoxon test, *, **, ***, **** for *p* < 0.05, 0.01, 0.001 and 0.00001 respectively. C-Comparison of overall survival estimated by the Kaplan–Meier method according to hierarchical clustering of CNV data (cluster 1 to cluster 4, log-rank test, p = 0.0001) D-Forest plot for CNV subgroup for overall survival (number of dead samples in each class: 39/41, 17/23, 8/8, 18/21). The hazard ratios (HR) and 95% confidence intervals (CI) were estimated by the Cox proportional hazard regression model. The Cox model was adjusted on EGFR (overexpression, negative, unknown), PTEN (PTEN loss, unknown) and treatment indicator (datasatinib, erlotinib and everolimus). Black box size is proportional to CI confidence= hazard ratio

### Chromosomal copy number abberations and their association with DIPG outcome

We then evaluated the impact of the CNA-based classification in four clusters (Fig. 2) on overall survival (OS) and showed that it was strongly correlated with OS (log rank test p=0.0001, Cox model; Fig. 4C). The median OS of C1 (39 deaths/41 patients), C2 (17/23), C3 (8/8) and C4 (18/21) were 0.71 [0.49-0.79], 1.23 [1.02-1.39], 1.24 [0.52-1.60] and 0.90 years [0.73-1.31], respectively. The hazard ratios were HR1/4= 1.96 [1.06-3.64], HR2/4=0.50 [0.24-1.03], HR3/4=0.60 [0.24-1.46] (Fig. 4D) (AIC=583.876 and Uno-c index=0.6869 (se=0.0347)). Alternative classifications based on the upcoming WHO classification of diffuse midline gliomas (H3.1^K27M^, H3.3^K27M^, H3^WT^)(Supplementary Fig. 5A) or a composite classification based on H3.1-K27M, H3.3-K27M, *TP53* and *PPM1D* mutations did not predict outcome of the cohort more precisely based on the AIC index (AIC 586.824 and 586.669 respectively, Supplementary Fig 5B). As patient stratification based on hierarchical clustering cannot be used easily in a prospective way, and because the computed chromothripsis score (30) could not reliably classify these patients (*data not shown*), it would desirable to establish a classification taking into account simplified CNA information. The combination of double mutated status of H3.3/TP53 in conjonction with chromosome 1q and chromosome 2 gains information can be used as surrogate for the CNA-based classification (Supplementary Fig. S4D).

### Mutational processes in DIPG

Mutational signature analysis showed a major contribution of signatures #3 (associated with DNA double-strand break-repair by homologous recombination failure and linked to a BRCAness phenotype)(31), #1 (age-association) and to a lesser extent to signatures #13 (APOBEC mutation signature previously linked to chromothripsis and *TP53* mutations)(13), #15 (defective DNA mismatch repair) and #22 (aristolochic exposure, Supplementary Fig. S6A). Signatures #3 and #13 were described to be linked with genomic instability which is in line with CNA signatures (13,31). We did not observe major differences of the signature contributions with respect to the CNA clusters (Supplementary Fig. S6B). The signature contribution was different from the one published on the subset of the ten-most frequently mutated HGG-K27M patients showing a higher contribution of signatures #8 and #16 while signature #15 and #22 were absent (13). Only four patients in our cohort (4.2%) harbored a mutational signature profile close to the HGG-K27M presented in Gröbner *and coll*., with very small contribution or absence of signature #3, which was predominant in the remaining samples of our cohort(13). However, these four samples did not show obvious common mutations and were distributed in the different CNA clusters.

### Overall portrait of recurrent SNV

Analysis of somatic single nucleotide variations (SNV) refined the frequency of the main DIPG alterations (Fig 5A & Supplementary Fig. S7A, C). Seventy-five percent of patients harbored H3.3-K27M mutation and among them around 2/3 were *TP53*^*MUT*^ (n=46/71). Eighteen patients were H3.1-K27M (19%) including fourteen (77.8%) presenting an associated *ACVR1* mutation and one sample (5.6%) with *TP53* mutation. Six patients were H3-wild-type and among them five were *ACVR1*-mutated and EZHIP positive in accordance with our previous publication (15) and one showed a H3K27me3 loss by IHC.

**Figure 5:**
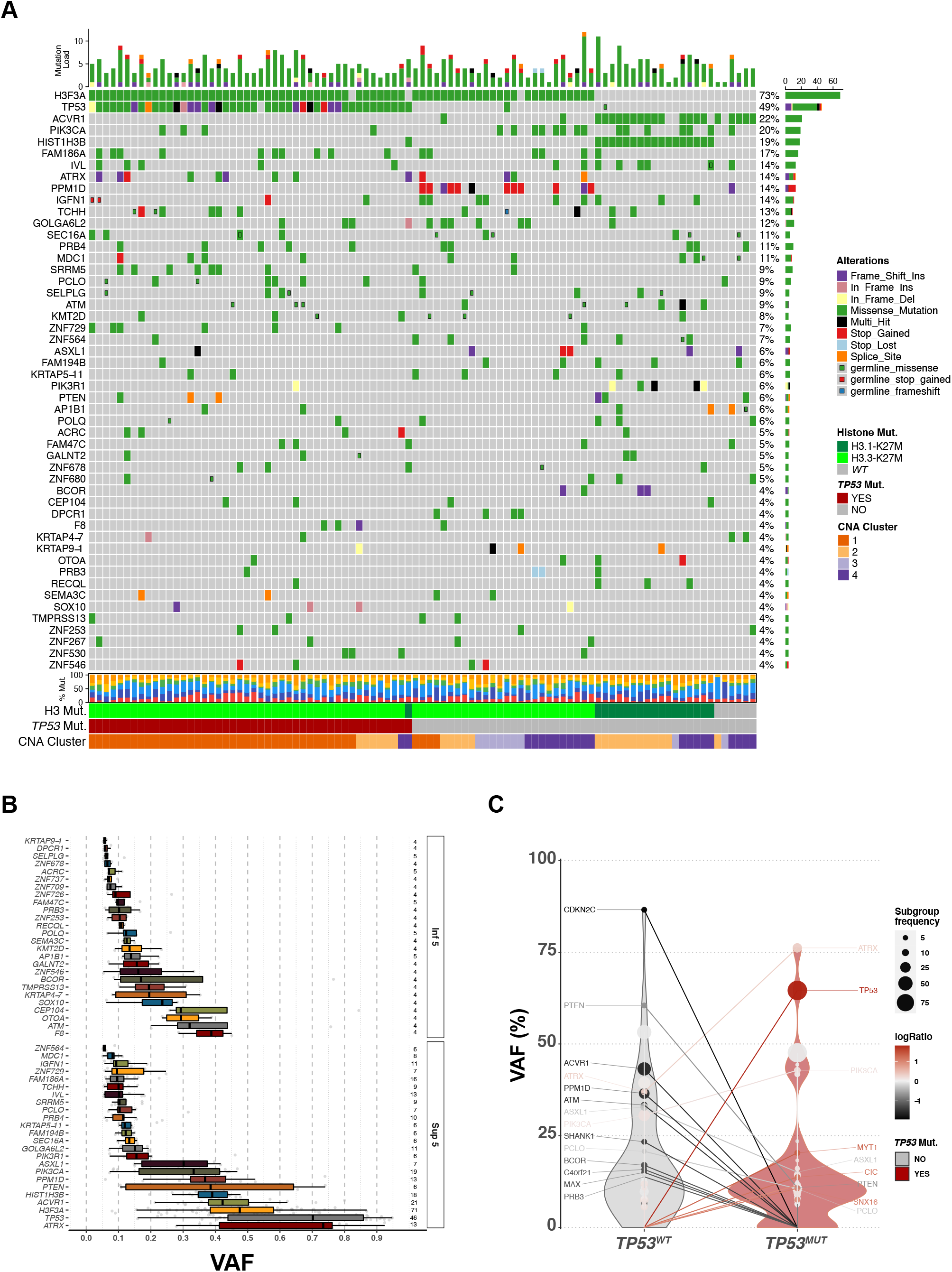
Summary of recurrent alterations in DIPG. A-Oncoplot depicting the top 50 mutated genes sorted and ordered by decreasing frequency. The top barplot shows the total number of somatic non-synonymous coding mutations for each patient, while the right barplot indicates the frequency of mutations in each gene taking into consideration the union of Mutect and Varscan callings. Only somatic SNV with more than 5 supporting reads associated and a VAF exceeding 5% in both Mutect or Varscan calling are shown on the plot. Germinal alterations with more than 5 supporting reads associated and a VAF exceeding 40% are indicated as a small square. Two patients harbor a H3.3K27M alteration with only 5 supporting reads and a sequencing depth of 44 and 49 for this gene and are indicated below as mutated for their H3 mutational status (asterisk). Tumors were grouped according to *TP53* mutational status. The repartition of transversion and transition per patient is shown in the barplot below the heatmap. B-Distribution of the VAF for the top 50 mutated genes. The number of patients presenting on alteration is indicated on the right. C-Violin plot of the mutated genes for which the VAF vary significantly between *TP53*^*MUT*^ and *TP53*^*WT*^ tumors. Only genes presenting a difference in median VAF exceeding 10 % between the 2 subgroups and mutated in at least 3 patients in one subgroup (with a VAF >5%) are reported. The median VAF per gene is shown with the number of mutated patients in each condition, either *TP53*^*WT*^ or TP53 mutated, indicated as the frequency and colored in red if the VAF is higher in *TP53*^*MUT*^ subgroup and in gray otherwise.

Nine already known recurrent alterations were found in more than 5 patients with a median VAF higher than 20% (Fig. 5B). Among them *PTEN* and *ASXL1* alterations are found in only 6 and 7 distinct samples respectively, with a huge range of VAF for *PTEN*. The other mutations *(i*.*e. in ACVR1, ATRX, H3F3A, HIST1H3B, PI3KCA, PPM1D* and *TP53*) were found in more than 13 patients. Fifteen additional genes were found mutated in at least 6 patients but are associated with lower range of VAF, likely reflecting later subclonal alterations. Among the most recurrent alterations, we identified few unrecognized mutations in *FAM186A* found mostly in H3.3 mutated tumors (15/16), *GOLGA6L2* (n=11) and *IGFN1* (n=11) both frequently found in conjunction with *PPM1D* alterations (n=4/11 and p=0.04), *IVL* (n=13) co-segregating with *BCOR* (3/4, p=0.008) and to a lesser extent with *PIK3CA (5/19* p=0.02) (Supplementary Fig. S7C&E).

*PIK3CA* alteration are enriched in *TP53*^*WT*^ (15/19 *TP53*^*WT*^ and 4/19 *TP53*^*MUT*^, p=0.009) and co-occurred with *AP1B1* (p=0.001). *PIK3R1* alterations were mutually exclusive with H3.3-K27M (p= 0.005) and co-segregated with H3.1-K27M (p= 0.0008) and *ACVR1* mutations (p=0.001). Lastly, we observed a significant co-segregation of *PRB4* and *MDC1* alterations (p=0.004). The genes mutated in 5 patients or less in the cohort are globally associated with lower median VAF except for *ATM, F8, OTOA, CEP104* and *SOX10* with a range of VAF over 20% (Fig. 5B).

Beside preferential association of mutations, we looked for significant differences in VAF according to H3.3-K27M *vs*. H3.1-K27M mutations or *TP53*^*MUT*^ *vs. TP53*^*WT*^ (Fig. 5C & Supplementary Fig. S7D) for the genes found mutated in at least 3 distinct samples of one of these subgroups. A relatively higher variant allele frequency of *ATRX* and *PIK3CA* alterations were observed in *TP53*^*MUT*^ tumors suggesting their early occurrence during carcinogenesis, as CNA could not solely explain these high VAF. Specific alterations in this *TP53*^*MUT*^ subgroup affecting *CIC, MYT1* and *SNX16* were found in only three patients.

In *TP53*^*WT*^ samples, higher VAF were found in *ASXL1, PPM1D, PIK3R1, PTEN* and *POLQ*. Mutations in *CDKN2C* (n=3), *ATM* (n=4), *BCOR* (n=4) and *SHANK1* (n=3) appeared restricted to the *TP53*^*WT*^ subgroup, although in a small number of patients.

The genes among the 50 most frequently mutated ones which are also submitted to CNA in a subset of patients were mainly affected by one kind of copy number variation. Indeed, genomic gains were invariably encountered in patients with somatic alteration of *ACVR1* (n=13), *FAM186A* (n=4), *GALNT2* (n=3), *IGFN1* (n=8), *IVL* (n=8), *LRP1B* (n=2), *PIK3CA* (n=2), *PPM1D* (n=3), *TCHH* (n=4), *ZNF253* (n=2), *ZNF678* (n=3), whereas genomic losses were observed concomitantly with mutations in *AP1B1* (n=7), *CIC* (n=2), *NF1* (n=2), *PTEN* (n=4), *PRDM9* (n=2), *TP53* (n=24), *ZNF530* (n=2) (Supplementary Fig. S8B).

Missense variations accounted for the vast majority of SNV detected and indels appeared quite rare in the cohort even if enriched in the alterations of some genes like *ASXL1, ATRX, BCOR*, or *PIK3R1* (Supplementary Fig. S1B and S7B).

Only rare germline variations were detected in this selection of 50 genes (Fig. 5A), except for some genes for which the proportion of germline mutations was markedly higher *i*.*e. ATM* (5/9), *SEC16A* (5/10), *SELPLG* (4/9), *KMT2D* (4/8). Outside of the top 50 genes most frequently mutated somatically, germline alterations of 13 genes (*ALPK3, ENOSF1, EYS, FGFR4, GCKR, GRM6, HHLA1, KCNA10, SETBP1, SIGLEC1, SMAP1, SPTA1, ZFHX3*) were found in at least 3 patients of the entire cohort while the frequency in the healthy population was very low (gnomAD <1/10000).

### Recurrent somatic alteration in specific biological pathways

Pathway analysis can uncover candidate genes that would otherwise remain undetected in gene-based analyses. We first performed unbiased analysis using genes mutated in more than one patient (n=264) that revealed a significant enrichment of five GO biological processes: *‘cell adhesion’, ‘cell morphogenesis’, ‘developmental process involved in reproduction’, ‘embryo development’*, and *‘regulation of neurogenesis signaling pathways’* (Supplementary Fig. S8A; adjusted FDR < 6.65e-04). This reflects the link of DIPG biology with the deregulation of a normal neuro-developmental process and the alteration of tumor-microenvironment interactions that enables tumor formation (32). Interestingly, *NLGN3* encoding a secreted protein that increases glioma cell proliferation in an activity-dependent manner (33), was found mutated in 2 patients. Many other genes belonging to these five biological processes are also found mutated in one unique patient of the cohort, increasing the number of genes affected of about 2-3 times in a given pathway and their enrichment (*data not shown*).

We next examined signalling pathways comprising genes frequently reported as altered in DMG and reported both recurrent SNV and CNA in our cohort (Supplementary Fig. S8B). In total, 87/95 cases (91.6%) were found to harbour mutations in one or more of these 6 key biological processes.

Genome integrity pathway was predominantly affected by SNV in 73% of the entire cohort with mutual exclusivity of *TP53, PPM1D* and *ATM* mutations (Supplementary Fig. S7B). Somatic *BRCA2* deletion in 39 % (n=14) of samples within cluster C1, and a deleterious germline alteration was observed in one additional patient. *TP53BP1* loss was also quite frequent in this subset of samples.

PI3K-AKT-MTOR signalling pathway was also frequently targeted in DIPG with 37% of patients harbouring non-synonymous somatic SNV, mainly in *PIK3CA* (20%), *PIK3R1* (6%) and *PTEN* (6%) found in distinct samples. When considering also CNA, alteration of this pathway reached 92% of the cohort. Twenty patients were associated with loss of chromosomal material of *PTEN* genomic region and among them 4 also harboured a somatic mutation of *PTEN*. Among the eight samples without alteration of PI3K-AKT-MTOR signalling at the molecular level, PTEN loss was detected by IHC in half of them and undetermined in the 4 remaining ones. Disruption of this pathway thus appears as an essential biological mechanism in DIPG initiation and progression expanding the initial finding of mTOR phosphorylation in the ten autopsy patients from Zarghooni *et al(34)*.

The proportion of SNV affecting TGF-β signaling reflected almost only *ACVR1* mutations (22%) in C2 and C4 clusters. CNA of both *ACVR1* and *ACVR2A* resulted mainly from whole chromosome 2 gains in C2 cluster. By contrast, *SMAD4*, located within chromosome 18q but outside the recurrent genomic regions identified by Gistic analysis, was frequently subjected to deletion. It was more frequently lost in C1 cluster without activating mutation of *ACVR1*. Interestingly, *DACH1* located on chromosome 13 and encoding a chromatin-associated protein which interacts especially with SMAD4, was frequently subjected to genomic loss in the same samples. *DACH1* is involved in developmental neural differentiation and was shown to regulate tumor-initiating activity of glioma cells and exert a tumor suppressor role in GBM (35) and more largely to negatively regulate the TGF-β pathway.

The chromatin regulator pathway was also affected by both single-nucleotide and copy-number changes (38% SNV only *vs*. total of 88%) largely driven by *ATRX* alterations (14%) preferentially found in C1 cluster or the mutual exclusive *ASXL1* alteration (7%) followed by *BCOR*, the histone H3K4 methyltransferase *KMT2D* and the bromodomain-containing protein *BRD7* a component of SWI/SNF chromatin remodeling complex. Co-segregation of *ASXL1, BCOR, KMT2D* and *BRD7* mutations with H3.3 or H3.1 mutated tumors previously reported were however not confirmed (1).

Only few DIPG (9%) carried single nucleotide variation in transcription factor (TF) encoding genes. Mutation of *CIC* a transcriptional repressor in the (RTK)/Ras/ERK signalling are usually seen in adult oligodendroglial tumors with 1p19q-co-deletion(36). *MAX* was found mutated in *TP53*^*WT*^ tumors (n=3/95) and frequently gained in *TP53*^*MUT*^ samples associated or not with *MYC*/*MYCN* gains. Two patients were mutated within *MED12* gene and one in *TCF12*.

## Discussion

DIPG can no longer be considered as an invariably fatal disease that could be safely diagnosed with imaging only in case of a short clinical history. Other diagnoses exist, as often than 7% here and could blur the evaluation of new therapeutics, reflecting the requirement for a central pathological review or methyloma analysis to avoid such sample misclassification.

Our study shows, for the first time prospectively in a large trial population, that the clinical outcome of the patients varies substantially and that discrepancies could be largely explained by the molecular characteristics of the tumor. Building upon biomarkers previously described in retrospective heterogeneous cohorts such as the type of histone H3 mutated at Lysine 27 (11,14,37) or the presence of a *TP53* mutation (38), we now link genomic structural alterations to prognosis. We identify using unsupervised clustering of CNAs the cluster of DIPG with the shortest median overall survival (8.5 months; cluster 1) as the one associated with complex structural variations, these patients being at a significantly higher risk of death any time post-randomization. The patients from this cluster 1 are older, and are characterized as well by H3.3^K27M^ and *TP53* mutations suggesting that CNA in the genome of these tumors is linked with these two mutations. In contrast, two other clusters are characterized by chromosome 1q gain and frequent *ACVR1* mutations, with (cluster 2) or without (cluster 4) chromosome 2 gain. Patients from cluster 2 had a conversely better survival (median OS of 15 months) suggesting that chromosome 2 gain is a paradoxically favourable prognostic factor. Interestingly, with respect to the histone status, no differences were observed within C2 and C4 in the distribution of the three main type of alterations, *i*.*e. H3F3A, HIST1H3B/C* or wild-type Histone H3 together with *EZHIP* overexpression (15). The last cluster showed overall maintenance of genome integrity. We have already shown in the past that a biomarker driven model based on the type of histone H3 mutated at Lysine 27 predicted overall survival better than the available models based on clinical history and imaging data (39). We now suggest that a model taking into account CNA could predict outcome more accurately than mutation-based prognostic classifications.

As chromosomal instability seemed to be the hallmark of the most severe form of DIPG, we investigated the possible mechanisms for these alterations. Copy number alterations reflected double-strand DNA lesions and double-strand break repair problems. Mutational signature and CNA features signature analyses pointed towards chromothripsis and *TP53* mutations (31), increased in cluster 1, as well as DNA double strand breaks or BRCAness phenotype.

Our data differ from the report of Gröbner *et al*. (13) where *TP53* mutations were not shown to be associated with structural abnormalities and chromothripsis, in high-grade gliomas (as opposed to other pediatric cancers). Noteworthy, all DIPG presenting complex genome rearrangements were associated with a dysfunction in the TP53 pathway.

TP53 dysfunction is one of the most frequent alteration occurring in DIPG through different mechanism, *e*.*g*. mutation of *TP53* itself (40) or *PPM1D* truncating mutations (23) and we described here a new mechanism with the frequent loss of tumor-suppressor *TP53BP1* in 28% of the cluster 1 samples while this alteration was not seen in the other DIPG clusters. We can wonder if TP53 dysfunction should be considered instead of *TP53* mutation in the patient stratification; however, the small number of samples with either *PPM1D* mutation, *TP53BP1* loss or other alterations of TP53 pathway did not allow us to evaluate this alternative. *TP53BP1* has also TP53-independent functions as a key mediator of the DNA-damage response and a direct modulator of DNA double-strand break repair (41).

Nevertheless, all mutations affecting the TP53 pathway are not invariably associated with chromothripsis-like phenotype. Indeed, somatic and germline *ATM* alterations were scattered in all subgroups. Ataxia-Telangiectasia cancer predisposition syndrome has not been associated with a significant increased risk of glioma and germline mutation frequencies reported here have not been found in other neoplasms (42) except for breast cancer (43) and leukemia/lymphoma (44). Confirmatory studies are necessary to confirm the role of germline or somatic *ATM* mutation could play in the pathogenesis of some DIPG, especially those without *TP53* mutations. Interestingly, we and others have recently shown that *ATM* could represent a relevant target to increase DIPG radiosensitivity specifically in a *TP53*^*MUT*^ context (38,45).

It remains to be elucidated how chromosomal instability occurs in DIPG, but noteworthy only in H3.3^K27M^ tumors and not in H3.1^K27M^ tumors. While TP53 dysfunction is not inducing DNA damage by itself, it likely permits the survival of cells with DNA damage (46,47). Especially with respect to chromosome instability, TP53 function is needed to propagate whole chromosome segregation errors while TP53 deficiency allows the propagation of structural chromosome imbalances(48). When comparing the mutations spectrum in the different subtypes of DIPG, *ATRX* mutations which could play a role in chromosomal instability (49–51) were indeed significantly more frequent in *TP53*^*MUT*^ and H3.3^K27M^ tumors.

In the BIOMEDE trial, WES was performed to identify targetable alterations in case of failure of the first line treatment, assuming that DIPG did not diverge too much overtime. Indeed, the mutation landscape was shown to be very similar between diagnostic (10) and autopsy samples (11,12,52). The most frequent targetable gains/amplifications identified concerned *PDGFRA/KIT/KDR*, exclusively in clusters 1 and 2, *IGF1R* and *CDK4* in cluster 1, and *PARP1* in cluster 2. Low level gains of *MYC* were also observed in cluster 1 and 2. The *PDGFRA* amplicon was one of the first recurrent alteration described in DIPG (34,53). IGF1R has been shown as a potential target in gliomas (54–56), and CDK4 inhibition, recently tested in phase I for DIPG (57) may represent an option for the subset of patients with *CDK4* amplification. With respect to targetable mutations, previously known frequent alterations in *ACVR1, PPM1D, ATM*, and mTOR pathway were identified. The mTOR pathway was altered with mutations in 37% of cases and PTEN loss of protein expression was constant across the whole cohort. Concurrent PI3K/mTOR pathway alteration and *PTEN* deletion have been shown as oncogenesis drivers in some cancers (58,59) and more recently Mukherjee *and coll*. showed that coexistent *PTEN* loss and *PIK3CA* mutation hyperactivate mTOR pathway output (60). One can therefore consider that AKT-mTOR pathway activation through PTEN loss may be a prerequisite or at least a co-driver in DIPG oncogenesis as shown in some genetically-engineered mouse models (61,62). Consequently, these findings justify further exploration of this pathway as a relevant target in DIPG.

Finally, this preliminary report shows that molecular profiling of DIPG stereotaxic biopsies is feasible on a multi-institutional and international level prospectively in most of the samples and that deep intra-tumour trajectories are not necessary to achieve correct diagnosis which is important for the safety of this procedure.

In conclusion, WES molecular profiling at diagnosis of this DIPG cohort of unprecedented size was able to define more precisely the complexity of DIPG oncogenesis and enabled the construction of a more accurate prognostic score usable in the clinic. Copy-number analysis identified H3.3-K27M DIPG with TP53 dysfunction as the most aggressive groups, while other H3.1-, H3.3-K27M and H3-WT tumors were distributed among the 3 other groups. This model will now have to be tested in a prospective cohort.

The role of TP53 mutations in maintaining the growth of tumours with multiple chromosomal alterations is key for the most severe subset of DIPG while AKT-mTOR pathway increased activity is evidenced in all DIPG types and potentially targetable. Therapeutic targets can be identified by sequencing at diagnosis and pave the way for more personalized therapies in DIPG.

## Material and methods

### Patient cohort description

Human clinical samples and data were collected after receiving written informed consent in accordance with the Declaration of Helsinki and approval from the respective national and institutional review boards. Patients with brainstem tumors were enrolled in the BIOMEDE trial (NCT02233049) on the basis of the classical clinico-radiological characteristics: intrinsic brainstem tumors involving at least 50 percent of the pons with a short clinical history, *i*.*e*. less than 3 months between the first symptoms and diagnosis (7). A first informed consent was requested for performing a stereotactic biopsy to ascertain the histological diagnosis and conduct whole exome sequencing for the tumor as well as exome sequencing of the patient’s blood. In case the diagnosis was confirmed on the basis of a diffuse brainstem glioma presenting a H3K27me3 loss with or without H3K27M mutation (14), a second informed consent was requested to enter the randomized phase II trial. Only patients with confirmed histological diagnosis of DIPG entered the trial. Treatment allocation to one of the three drugs (erlotinib, everolimus and dasatinib) was based on the presence or absence of EGFR overexpression and the presence or absence of mTOR activation (PTEN loss of expression, pS6 and/or AKT overexpression) by immunohistochemistry. In case of the absence of the biomarker, the patient could not be randomized in the corresponding arm. As dasatinib has more than 60 targets, many of them being expressed in DIPG (17), no biomarker was requested to be randomized in this arm. Treatments were administered together with radiotherapy and in the adjuvant setting until progression or major toxicity.

### Tumor DNA preparation

Tumor DNA was extracted from frozen material using Allprep DNA/RNA and Blood DNA using Paxgene Blood DNA kits following manufacturer instructions (Qiagen, Germany). Purified DNA was quantified using the Qubit Broad Range double-stranded DNA assay (Life Technologies, Carlsbad, CA, USA).

### Immunohistochemistry and FISH analysis

All patients with initial local diagnosis of DIPG were reviewed centrally according to the WHO 2007 classification by a central BIOMEDE reference neuropathologist (PV) prior to enrollment. Four μM sections were stained by an automated Discovery XT or Benchmark XT (Ventana Medical Systems, Tucson, USA). The following primary antibodies were used: Ki-67 (1:200, clone MIB-1, Dako, Glostrup, Denmark), p53 (1:5000, clone DO-1, Santa Cruz Biotechnology, Dallas, USA), EGFR (prediluted, clone 3C6, Ventana, Burgess Hill, United Kingdom), PTEN (1:300, clone 6H2-1, Dako, Glostrup, Denmark), H3K27M (1:1000, clone ABE419, EMD Millipore, Billerica, USA), H3K27me3 (1:1250, clone C15410195, Diagenode, Seraing, Belgium), phospho-S6 (1:400, clone Ser235/236, Cell Signaling Technology, Danvers, USA) and phospho-AKT (1:100, clone 736E11, Cell Signaling Technology, Danvers, USA). Accumulation of TP53, EGFR score, and loss of PTEN were assessed as previously described(63,64). External positive and negative controls were used for all antibodies.

FISH study was performed on interphase nuclei according to the standard procedures and the manufacturer’s instructions. The copy number of the gene *PDGFRA* was assessed using the following centromeric and locus specific probe PDGFRA/CEN4p (Abnova, Taïwan).

Signals were scored in at least 100 non-overlapping intact interphase nuclei per case. Gene copy number per nucleus was recorded as follows: one copy, two copies, copy number gain (3), and amplification (≥4 copies or innumerable clusters). Copy gain/amplification were considered if they were detected in more than 10% nuclei. Results were recorded using a DM600 imaging fluorescence microscope (Leica Biosystems, Richmond, IL) fitted with appropriate filters, a CCD camera, and digital imaging software from Leica (Cytovision, v7.4).

### Survival analysis

Overall survival (OS, main endpoint of the BIOMEDE trial), defined as the time from the randomization date to death whatever the cause of death or from the date of last news for alive patients, was estimated by Kaplan-Meier method. For each prognostic classification scheme, OS was compared by the log-rank test. The association between prognostic classification schemes and OS will be quantified through the hazard ratio (HR) and its 95% confidence interval (CI) estimated by the Cox proportional hazard regression model. The Cox model will be adjusted on EGFR (overexpression, negative, unknown), PTEN (Pten loss, unknown) and treatment indicator (datasatinib, erlotinib and everolimus). These two markers are used for patient stratification in the BIOMEDE trial. The goodness-of-fit and discriminant ability of the different prognostic classification schemes will be compared by (i) the Akaike criterion (AIC) since the different prognostic classification schemes are not nested (lower AIC indicates better fit) and (ii) the Uno’s concordance statistic (c-index close to 1 indicates good discriminant ability), respectively. Standard error (se) of Uno’s c-index is calculated using 5000 perturbation samples. The cut-off date was January, 01 2019. The SAS software 9.4 was used for the survival analyses.

### Whole exome sequencing

Library preparation, exome capture and sequencing were performed by IntegraGen SA (Evry, France). Briefly, libraries were prepared from 150 ng of fragmented genomic DNA using the NEBNext Ultra DNA Library Prep Kit for Illumina (New-England Biolabs) and sequences captured using the SureSelectHuman clinical Research Exome V1 & 2 kit (Agilent) followed by paired-end 75 bases massively parallel sequencing on Illumina NextSeq 500 with a mean coverage for normal and blood 60 and 110X respectively. After base calling using the Real-Time Analysis (RTA2) software sequence pipeline and quality check by FastQC (v11.0.3), and fastqscreen, reads were mapped to the human genome build (GRCh37) using the Burrows-Wheeler Aligner (BWA mem) tool. Filtering and recalibration were conducted with picard tools following best practice in GATK v3.6.

### Somatic variant calling and Mutational signature

Somatic mutations including SBS (single base substitutions) and indels were detected by Mutect2 or Varscan (v2.3.9) and false positive filtered using Varscan fpFilter. Variants were then annotated by Variant Effected Predictor (v103) (65) and further converted to MAF files to be analysed by Maftools(66) using dbSNP version 153. We selected somatic variations that passed all quality control filters and associated with a sequencing depth in both normal and tumor samples higher than 10, a variant allele frequency (VAF) > 5 % and a minimum of 5 supporting reads in tumor and a VAF < 3 % and a maximum of 3 supporting reads in the matching normal samples. We filter out intronic, UTR and synonymous variations as well as single nucleotide variations (SNV) with a frequency higher to 1% in healthy donors from 1000 Genomes Project (1000G_phase3) and gNOMAD 2.1.

Mutational signature analysis was performed with SigMiner package following general pipeline. Mutational similarity with Cosmic v2(67) was estimated using sig.fit function in Sigminer package. Somatic interaction analysis (mutual exclusivity or co-occurence) was performed using pairwise Fisher exact test.

### SNV Germline analysis

Raw germline variants were initially filtered by removing those with a VAF below 40% and synonymous variations yielding to a median of 67 per sample (ranging from 27 to 110). Recurrent germline variants were selected by filtering SNV associated with a frequency below 1/10000 in gNOMAD and shared by at least 3 distinct patients (median number of mutations: 9, ranging from 1 to 53).

### Copy Number Analysis and CN signature

Copy number variations were investigated using EaCoN (R packages: https://github.com/gustaveroussy/EaCoN) with FACETS segmentation tool. Clustering analysis and visualization were performed with R packages tidyverse, pheatmap. Pairwise similarity of samples was calculated using Euclidean distance and agglomeration performed using Ward’s linkage method. Minimal common regions of alterations were retrieved based on focality, amplitude, and recurrence of alterations by Gistic2 (genomic identification of significant targets in cancer) analysis.

Samples presenting a log2ratio higher than 0.20 for at least 60% of the 1q chromosome arm or the entire chromosome 2 were labelled as “chrs 1q gain” and “chrs2 gain” respectively for subsequent analysis. Amplification of genomic region was defined as associated with a log2 ratio>1.5.

CN signature were performed using SigMiner package following general pipeline as described in (30). We use non-negative matrix factorization (NMF) to deconvoluate a matrix of 8 component from copy number profile. Estimation of the best number of signatures was optimized with looking silhouette profile and cophenetic metric and we select n = 3. Chromothripsis score was computed as already published(68) and calculated as follow: based on the presence of ten to hundreds of locally clustered segmental losses being interspersed with regions without copy number, *CS =* Σ_*Chr*_ *N*^*2*^*Oscn* where N is square number of patterns “2-1-2” copy number alteration.

The chromothripsis score was computed as previously published (30).

## Data Availability

Data are currently being deposited on EGA (https://ega-archive.org). Please contact authors for updates.

## Acknowledgments

The authors would like to thank the parents and patients who participated in the study and the investigators who treated the patients in the BIOMEDE trial. We acknowledge also the support of the Institut National du Cancer (Programme Hospitalier de Recherche Clinique), the charities Imagine for Margo (to TK and JG), l’Etoile de Martin (to DC) and the Lemos Family (to JG).

## Notes

### Competing Interest Statement

The authors have declared no competing interest.

### Clinical Trial

NCT02233049

### Author Declarations

The IRB 'Comite de Protection des Personnes d'Ile de France' provided an approval on July 22nd 2014 for the research within the BIOMEDE clinical trial under the file # 2014-001929-32. The French oversight body 'Agence Nationale de Securite du medicament et des produits de sante' (ANSM) delivered an autorization for the clinical trial and associated biomedical research under the reference 140670A-12 on August 6th 2014.

