## Supplemental Figure S1 to S8 for "Copy-number alterations reshape the classification of diffuse intrinsic pontine gliomas. First exome sequencing results of the BIOMEDE trial"

**Supplemental Figures S1 to S8**

**---**

### Copy-number variations reshape the classification of diffuse intrinsic pontine gliomas. First exome sequencing results of the BIOMEDE trial.

Thomas Kergrohen[1,2], David Castel[1,2], Gwenael Le Teuff[3], Arnault Tauziède-Espariat[4], Emmanuèle Lechapt-Zalcman[4], Karsten Nysom[5], Klas Blomgren[6,7], Pierre Leblond[8], Anne-Isabelle Bertozzi[9], Emilie De Carli[10], Cécile Faure-Conter[11], Celine Chappe[12], Natacha Entz-Werlé[13], Angokai Moussa[3], Samia Ghermaoui[1], Emilie Barret[1], Stephanie Picot[1], Marjorie Sabourin-Cousin[1], Kevin Beccaria[1,14], Gilles Vassal[1,2], Pascale Varlet[4], Stephanie Puget[14], Jacques Grill[1,2] and Marie-Anne Debily[1,15].

1 Molecular Predictors and New Targets in Oncology, INSERM, Gustave Roussy, Université Paris-Saclay, Villejuif, France.
2 Département de Cancérologie de l'Enfant et de l'Adolescent, Gustave Roussy, Université Paris-Saclay, Villejuif, France.
3 Département de Biostatistique et d'Epidémiologie, Gustave Roussy et INSERM U1018 CESP, Université Paris-Sud, Université Paris-Saclay, Equipe OncoStat, Villejuif, France.
4 Department of Neuropathology, GHU Paris-Neurosciences, Sainte-Anne Hospital, Paris, France.
5 Department of Pediatrics and Adolescent Medicine, Rigshospitalet, Copenhagen, Denmark.
6 Department of Pediatric Hematology and Oncology, Karolinska Institutet, Stockholm, Sweden.
7 Department of Pediatric Hematology and Oncology, Karolinska University Hospital, Stockholm, Sweden.
8 Département d'oncologie pédiatrique, Centre Oscar Lambret, Lille, France (PL).
9 Département d'Hématologie et d'Oncologie Pédiatrique, Hôpital Purpan, Centre Hospitalier Universitaire, Toulouse, France.
10 Département d'Hématologie et d'Oncologie Pédiatrique, Centre Hospitalier Universitaire, Angers, France.
11 Institut d'Hématologie et d'Oncologie Pédiatrique de Lyon (IHOPE), Lyon, France.
12 Département d'Hématologie et d'Oncologie Pédiatrique, Centre Hospitalier Universitaire, Rennes, France.
13 UMR CNRS 7021, Laboratory Bioimaging and Pathologies, Tumoral signaling and therapeutic targets, Faculty of Pharmacy, Illkirch, and Pediatric Onco-hematology unit, University Hospital of Strasbourg, France.
14 Département de Neurochirurgie Pédiatrique, Hôpital Necker Enfants Malades, Paris, France.
15 Département de Biologie, Univ. Evry, Université Paris-Saclay, Evry, France.

Corresponding authors

Dr Marie-Anne Debily and Dr Jacques Grill
U981, Prédicteurs Moléculaires et Nouvelles Cibles en Oncologie, INSERM, Gustave Roussy, Université Paris-Saclay, Équipe "Génomique et Oncogénèse des Tumeurs Cérébrales Pédiatriques', Bâtiment de Médecine Moléculaire,
114 rue Édouard Vaillant, 94805 Villejuif cedex, France

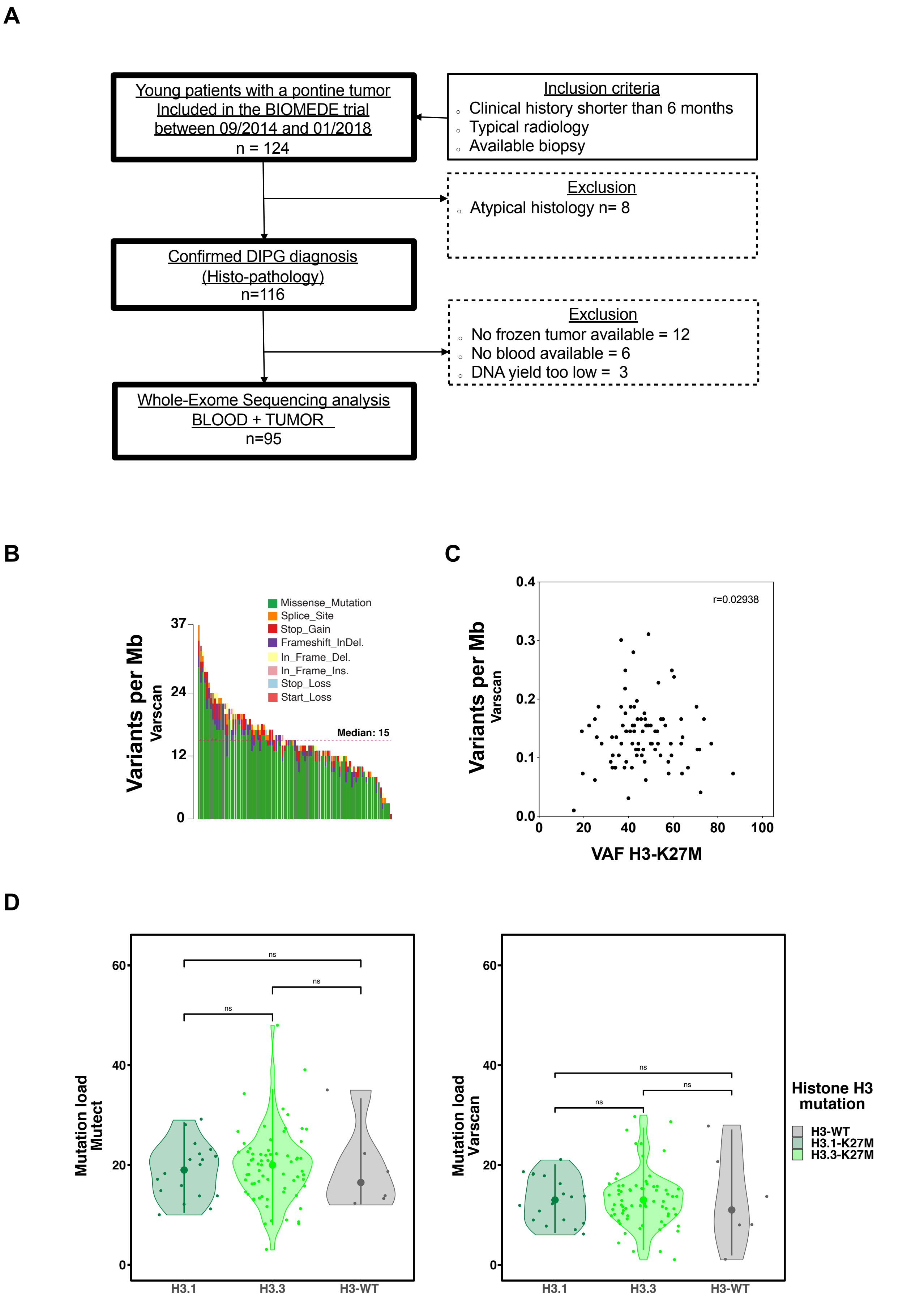
A
Young patients with a pontine tumor
Included in the BIOMEDE trial
between 09/2014 and 01/2018
n = 124
Inclusion criteria
Clinical history shorter than 6 months
Typical radiology
Available biopsy
Exclusion
Atypical histology n= 8
Confirmed DIPG diagnosis
(Histo-pathology)
n=116
Exclusion
No frozen tumor available = 12
No blood available = 6
DNA yield too low = 3
Whole-Exome Sequencing analysis
BLOOD + TUMOR
n=95
B
Variants per Mb
Varscan
Missense_Mutation
Splice_Site
Stop_Gain
Frameshift_InDel.
In_Frame_Del.
In_Frame_Ins.
Stop_Loss
Start_Loss
Median: 15
C
Variants per Mb
Varscan
r=0.02938
VAF H3-K27M
D
Mutation load
Mutect
Mutation load
Varscan
ns
H3.1
H3.3
H3-WT
Histone H3
mutation
H3-WT
H3.1-K27M
H3.3-K27M

**Supplemental figure S1**

**A-** Flow chart of DIPG patients in the present study including the inclusion and exclusion criteria. **B-** Stacked barplot representing the number and types of variants per sample using Varscan variant caller. **C-** Variants per Mb detected according to the VAF of the H3-K27M mutation for all H3 mutated tumors using VarScan variant caller (Spearman correlation 0.02938, $p$ = 0.7883). **D-**Violin plots representing the number of variants per Mb for H3.3-, H3.1-K27M as well as H3-WT patients using either Mutect (left panel) or VarScan (right panel) variant callers. No significant difference was found between the 3 subgroups (Wilcoxon test).

Figure S2

A

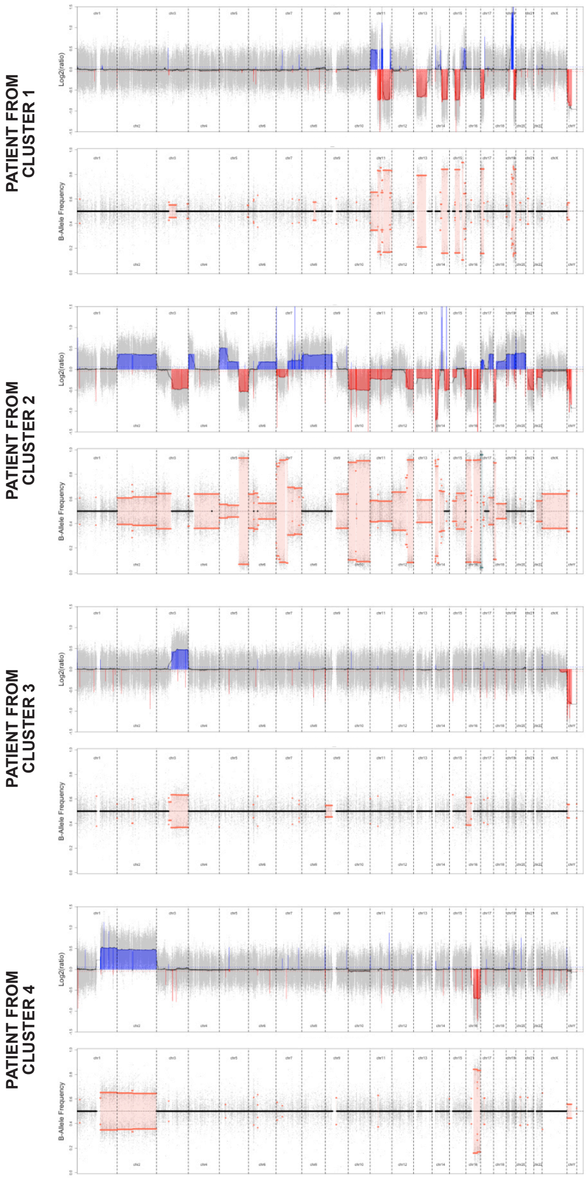
PATIENT FROM CLUSTER 1
PATIENT FROM CLUSTER 2
PATIENT FROM CLUSTER 3
PATIENT FROM CLUSTER 4
Log2(ratio)
B-Allele Frequency

B

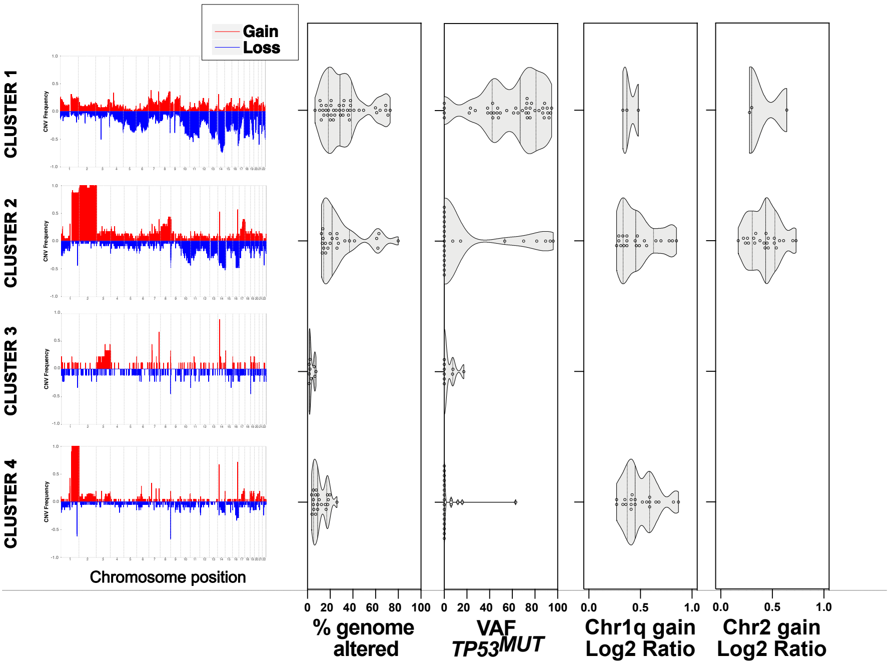
Gain
Loss
CLUSTER 1
CLUSTER 2
CLUSTER 3
CLUSTER 4
CNV Frequency
Chromosome position
0 20 40 60 80 100
% genome altered
0 20 40 60 80 100
VAF TP53MUT
0.0 0.5 1.0
Chr1q gain Log2 Ratio
0.0 0.5 1.0
Chr2 gain Log2 Ratio

Figure S2

**C**

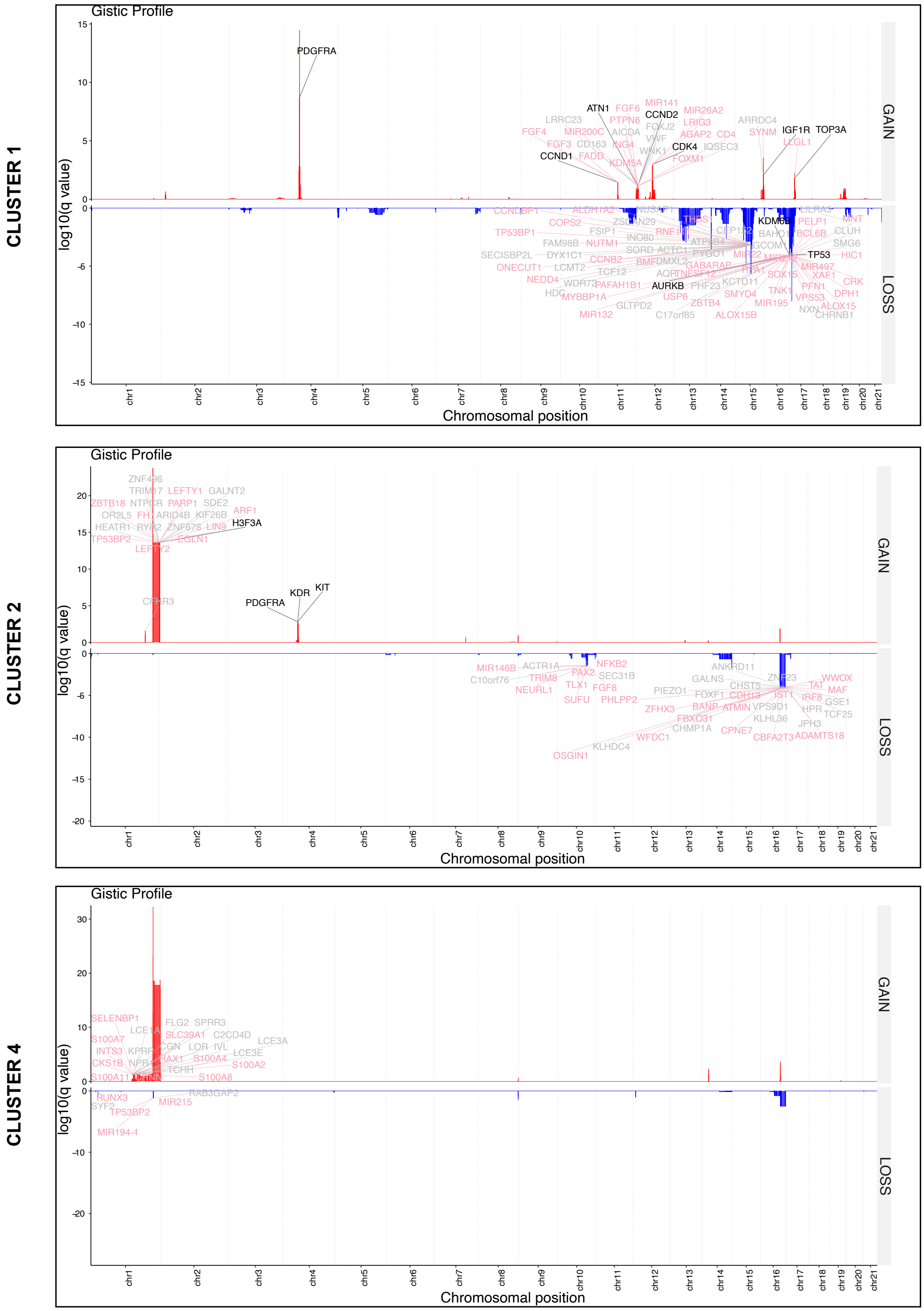
CLUSTER 1
Gistic Profile
log10(q value)
GAIN
LOSS
PDGFRA
ATN1
CCND2
CDK4
CCND1
IGF1R
TOP3A
KDM6B
TP53
AURKB
Chromosomal position
CLUSTER 2
Gistic Profile
log10(q value)
GAIN
LOSS
H3F3A
KDR
KIT
PDGFRA
Chromosomal position
CLUSTER 4
Gistic Profile
log10(q value)
GAIN
LOSS
Chromosomal position

**Supplemental figure S2**

**A**-Representative CNV profiles from each of the 4 main clusters from figure 2.
The genomic copy number aberrations (CNA) presented were computed by comparing the tumor exome data to the normal blood exome data. On the top panel, the log2ratio signal is plotted against chromosomal position, each dot representing a locus. The DNA segments resulting from the joint segmentation are colored in red and blue for gains and losses respectively for log2ratio higher than 0.25 or smaller than -0.25. The dotted horizontal lines represent a threshold of -0.05 and 0.05. On the bottom panel, the log2 B-allele frequency (BAF) signal is presented and labelled in orange for differences > 0.55.
**B**- On the left, average profile of CNV for each of the 4 clusters from figure 2. Frequency of copy-number gains (red) and losses (blue) are plotted along the genomic location. On the right, violin plots reflecting for each cluster the percentage of genome associated with CNA, the VAF of *TP53* mutations, chromosome 1q and chromosome 2 gains. Gain was defined as a log2 ratio of CNA > 0.25 for at least 80% of chromosome arm.
**C**- GISTIC analysis of copy number alterations for each DIPG CNV clusters. Significance is reported as false discovery rate-corrected q-value along the y axis (log10 q-value for gain and -log10 q-value for loss) with chromosomal position along the x axis. Genes previously reported as altered (with SNV or CNV) in DIPG from the study of MacKay *et al.(1)* are labeled in black, genes annotated as known oncogene or tumor suppressor genes are labeled in pink and those found mutated in our cohort by either Mutect or Varscan variant callers in gray. No recurrent CNA segment were identified for Cluster 3.

Figure S3

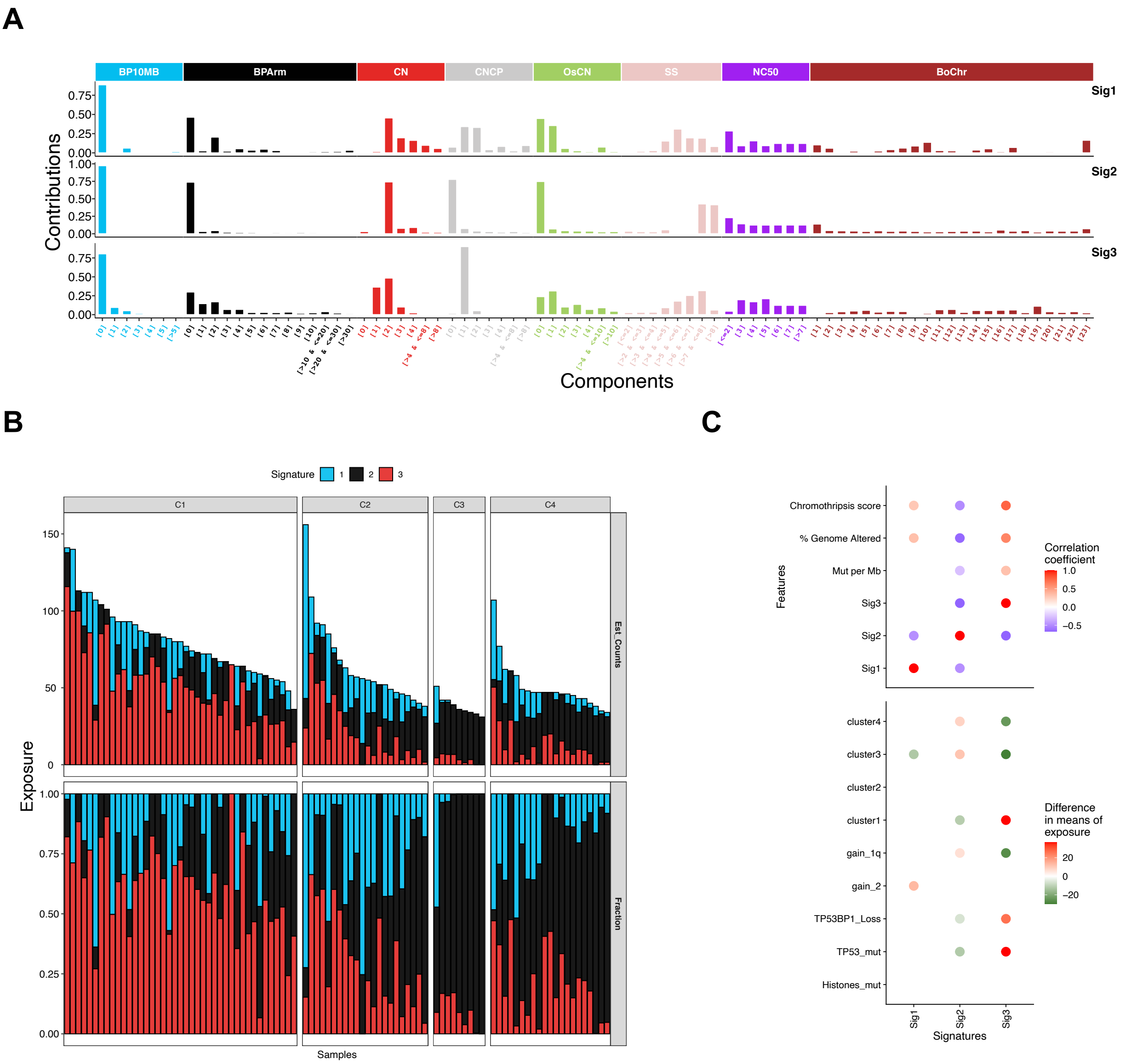
A
BP10MB
BPArm
CN
CNCP
OsCN
SS
NC50
BoChr
Sig1
Sig2
Sig3
Contributions
Components
B
Signature 1 2 3
C1
C2
C3
C4
Est_Counts
Fraction
Exposure
Samples
C
Chromothripsis score
% Genome Altered
Mut per Mb
Sig3
Sig2
Sig1
Features
Correlation coefficient
cluster4
cluster3
cluster2
cluster1
gain_1q
gain_2
TP53BP1_Loss
TP53_mut
Histones_mut
Difference in means of exposure
Signatures

**Supplemental figure S3**

A- Copy number signatures identified in DIPG from WES data. Copy number signature has 8 different features with 80 components and was row normalized within each feature; *i.e.* the breakpoint count per 10 Mb (BP10MB); the breakpoint count per chromosome arm (BPArm); the absolute copy number of the segments (CN); the difference in copy number values between adjacent segments (copy number change point, or CNCP); the lengths of oscillating copy number segment chains (OsCN); the log10 based size of segments (SS); the minimal number of chromosomes with 50% copy number alterations (NC50); the distribution of copy number alterations in each chromosome (burden of chromosome, named "BoChr").
B- For each patient estimated copy number segment counts (top panel) and the relative contribution (bottom panel) of each signature are shown as a staked barplot according to C1 to C4 clusters.
C- Associations between the exposures of genome alteration signatures or the difference of exposure and molecular features. Mutation per Mb was computed from Mutect variant calling. Pearson correlations for exposure and Kruskal Wallis for difference in mean of exposure with false-discovery rate $P < 0.05$ are color-coded.

Figure S4

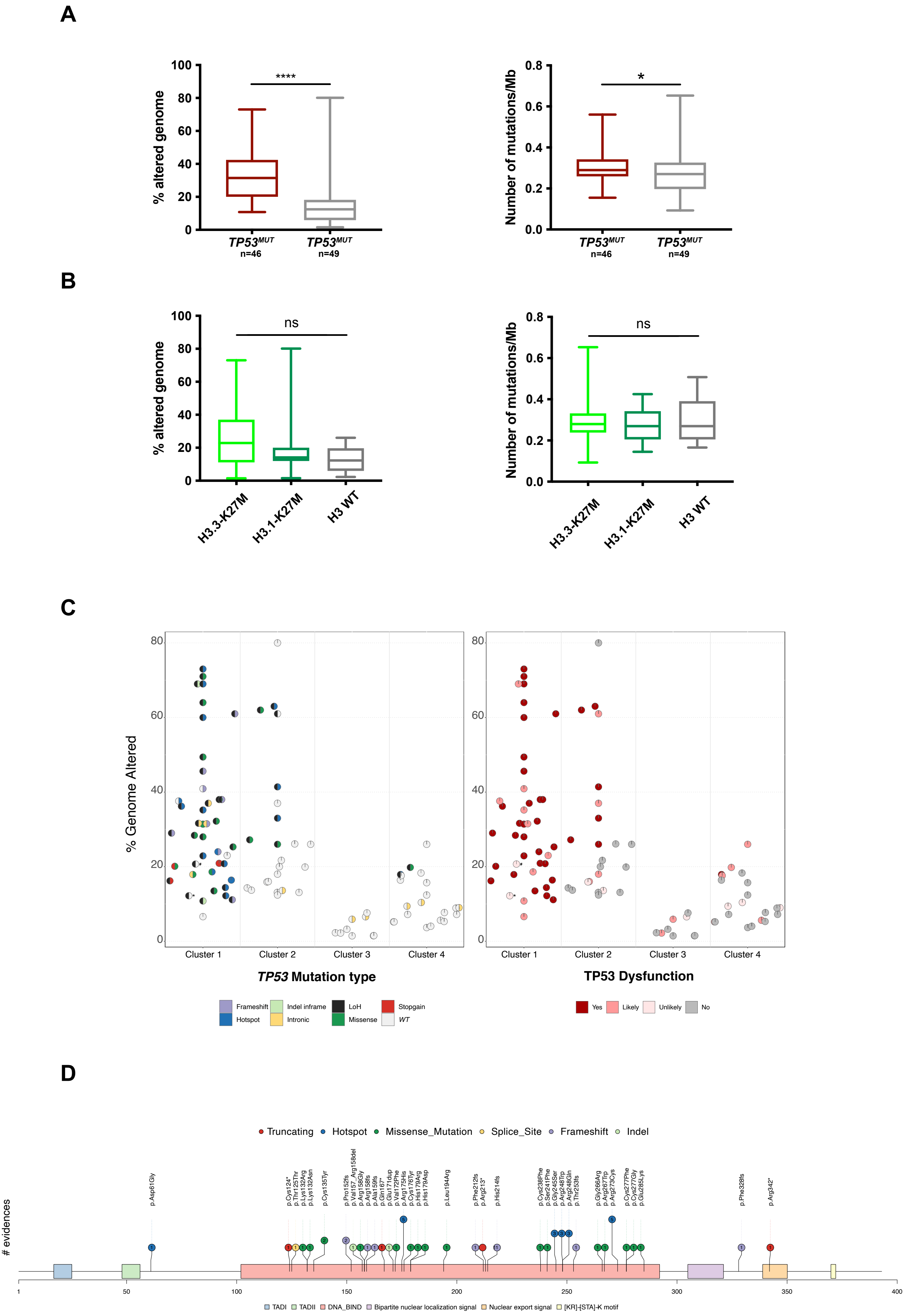
A
% altered genome
****
TP53^MUT n=46
TP53^MUT n=49
Number of mutations/Mb
*
B
ns
H3.3-K27M
H3.1-K27M
H3 WT
C
% Genome Altered
Cluster 1
Cluster 2
Cluster 3
Cluster 4
TP53 Mutation type
TP53 Dysfunction
Frameshift
Indel inframe
LoH
Stopgain
Hotspot
Intronic
Missense
WT
Yes
Likely
Unlikely
No
D
Truncating
Hotspot
Missense_Mutation
Splice_Site
Frameshift
Indel
### evidences
p.Asp61Gly
p.Cys124*
p.Thr125Thr
p.Lys132Arg
p.Lys132Asn
p.Cys135Tyr
p.Pro152fs
p.Val157_Arg158del
p.Arg158Gly
p.Arg158fs
p.Ala159fs
p.Gln167*
p.Glu171dup
p.Val172Phe
p.Arg175His
p.Cys176Tyr
p.His179Arg
p.His179Asp
p.Leu194Arg
p.Phe212fs
p.Arg213*
p.His214fs
p.Cys238Phe
p.Ser241Phe
p.Gly245Ser
p.Arg248Trp
p.Arg248Gln
p.Thr253fs
p.Gly266Arg
p.Arg267Trp
p.Arg273Cys
p.Cys277Phe
p.Cys277Gly
p.Glu285Lys
p.Phe328fs
p.Arg342*
TADI
TADII
DNA_BIND
Bipartite nuclear localization signal
Nuclear export signal
[KR]-[STA]-K motif

**Supplemental figure S4:**

Box plot reflecting the percentage of genome submitted to CNV according to *TP53* mutational status (A)(*TP53*$^{MUT}$ n=46 and *TP53*$^{WT}$ n=49, Wilcoxon test, *p*= 8.6e-10) or histone H3 mutational status (B)(H3.3-K27M n=71, H3.1-K27M n=18 and H3-WT n=6, Wilcoxon test, H3.3- *vs.* H3.1-K27M *p* =0.19, H3.3- *vs.* H3WT *p* =0.19, H3.1 *vs.* H3-WT *p* =0.46). Right panel: box plot representing the number of non-synonymous somatic SNV per Mb detected by Mutect caller according to *TP53* mutational status (Wilcoxon test, *p* = 0.0473, median of mutations/Mb about 0.29 for *TP53*$^{MUT}$ and 0.27 for *TP53*$^{WT}$) or histone H3 mutational status (H3.3- *vs.* H3.1-K27M *p* =0.439, H3.3- *vs.* H3WT *p* =0.909, H3.1 *vs.* H3WT *p* =0.738, median of mutations/Mb about 0.28 for H3.3-K27M, 0.27 for H3.1-K27M, 0.27 for H3-WT).
C- Scatter dot plots presenting the percentage of genome submitted to CNV per patient according to the type of variations (left panel) or the impact of the variation on TP53 functionality (right panel). Samples with 'Unlikely Pathogenicity' presenting *TP53BP1* loss or a somatic mutation of *NELFCD* are highlighted with the symbols * and #, respectively.
D – Lolliplot of *TP53* alterations in the cohort presenting the distribution of the variations along the amino acid sequence. The number of patients presenting each modification is indicated in the circles and the color-code of alterations indicated above the plot.

Figure S5

A

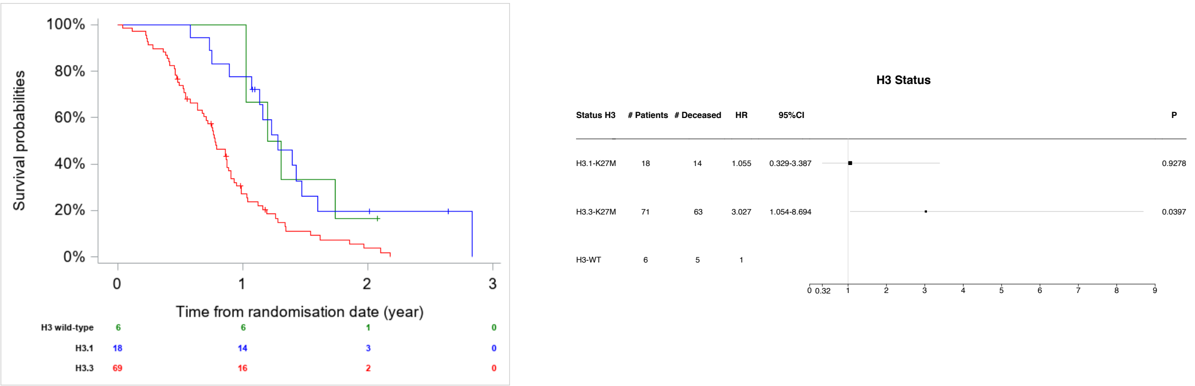


B

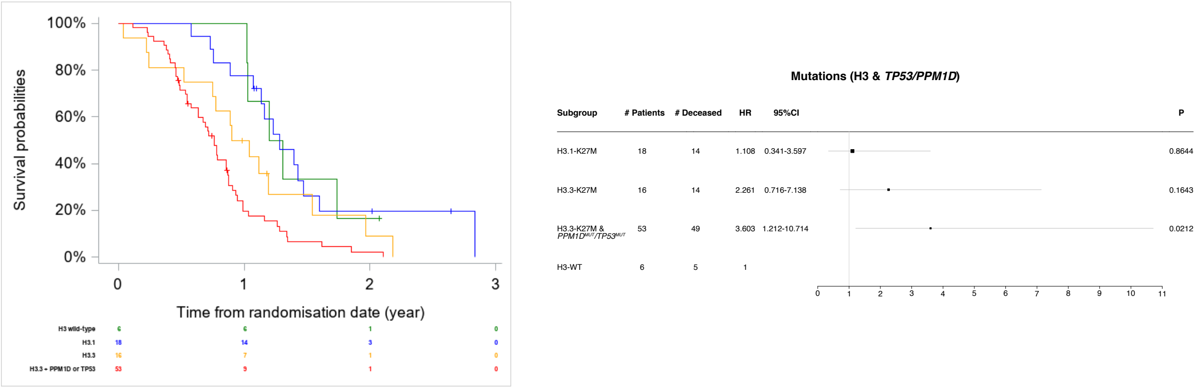


C

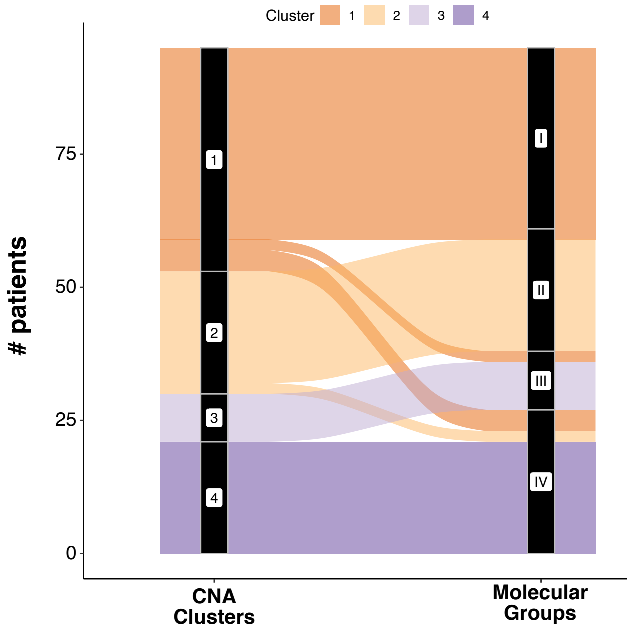

**Supplemental figure S5:**

A- Left panel: comparison of overall survival by using the Kaplan–Meier method according to hierarchical clustering of CNV data (cluster 1 to cluster 4, log-rank test, $p = 0.0001$) or H3 mutational status (H3.1-K27M, H3.3-K27M, H3.3-K27M $TP53^{MUT}$ and/or $PPM1D^{MUT}$ and H3-WT, log-rank test, $p = 0.0009$). Right panel: forest plot for overall survival of patients stratified by molecular alterations *i.e.* H3.1-K27M, H3.3-K27M, and H3-WT (number of dead samples at the time of the analysis in each class: 14/18, 63/69, 49/53, 5/6).

B- Left panel: comparison of overall survival by using the Kaplan–Meier method according to hierarchical clustering of CNV data (cluster 1 to cluster 4, log-rank test, $p = 0.0001$) or H3 mutational status (H3.1-K27M, H3.3-K27M, H3.3-K27M $TP53^{MUT}$ and/or $PPM1D^{MUT}$ and H3-WT, log-rank test, $p = 0.0009$). Right panel: forest plot for overall survival of patients stratified by molecular alterations *i.e.* H3.1-K27M, H3.3-K27M, H3.3-K27M $TP53^{MUT}$ and/or $PPM1D^{MUT}$ and H3-WT (number of dead samples at the time of the analysis in each class: 14/18, 14/16, 49/53, 5/6).

C- Riverplot showing distribution of DIPG samples among the CNA-based clusters on the left and molecular groups based on both chromosome 1q and 2 gains and H3.3K27M/TP53 mutations on the right following this classification: Group I, DIPG H3.3-K27M TP53MUT without 1q and 2 chromosomal imbalances; Group II, DIPG with chromosome 2 gain; Group III, DIPG with chromosome 1q gain and Group IV, DIPG TP53WT without 1q and 2 chromosomal imbalances.

### Figure S6

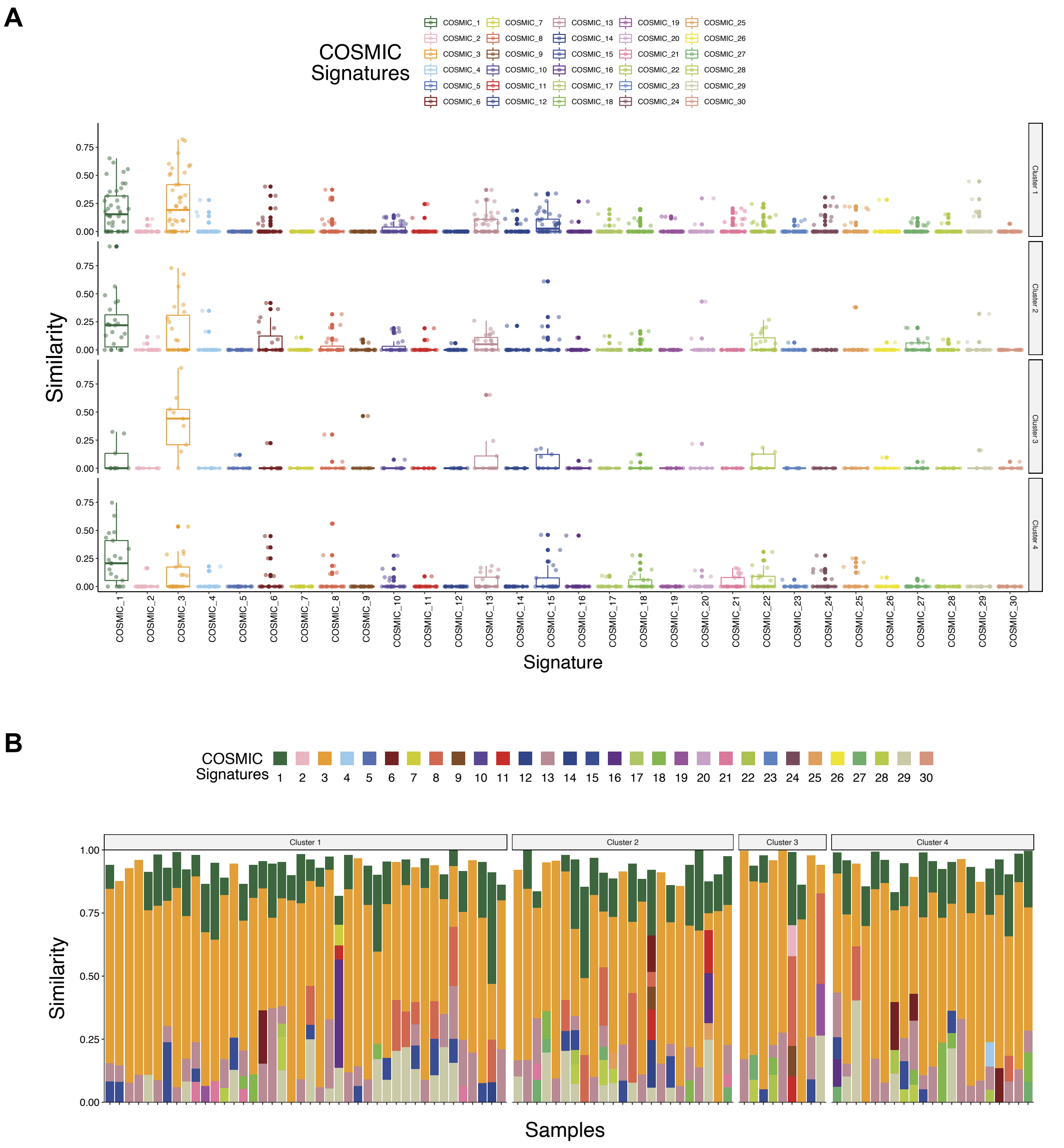
A
COSMIC Signatures
Similarity
Signature
Cluster 1
Cluster 2
Cluster 3
Cluster 4
B
COSMIC Signatures
Similarity
Samples

**Supplemental figure S6:**

**A-** Box plot presenting the similarity of the mutational profile of each patient to the thirty mutational signatures(69). Patients are subdivided according to CNA clusters.
**B-** Contributions of the thirty known mutational signatures to the somatic mutations; only signature with a contribution exceeding 5% are kept, each bar represents one individual tumor.

### Figure S7

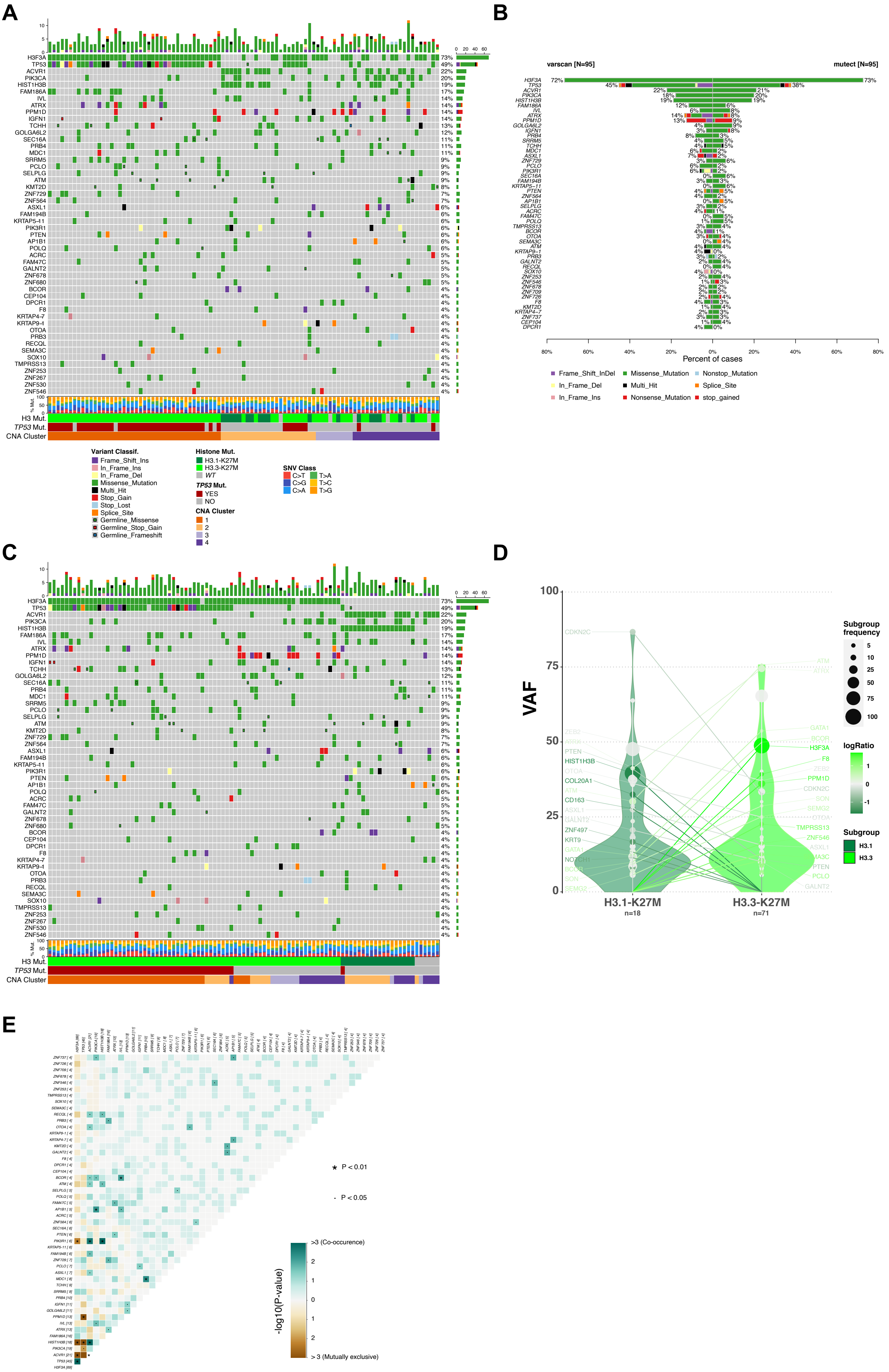

A
B
varscan [N=95]
mutect [N=95]
Percent of cases
Frame_Shift_InDel
Missense_Mutation
Nonstop_Mutation
In_Frame_Del
Multi_Hit
Splice_Site
In_Frame_Ins
Nonsense_Mutation
stop_gained
H3 Mut.
TP53 Mut.
CNA Cluster
Variant Classif.
Frame_Shift_Ins
In_Frame_Ins
In_Frame_Del
Missense_Mutation
Multi_Hit
Stop_Gain
Stop_Lost
Splice_Site
Germline_Missense
Germline_Stop_Gain
Germline_Frameshift
Histone Mut.
H3.1-K27M
H3.3-K27M
WT
TP53 Mut.
YES
NO
CNA Cluster
1
2
3
4
SNV Class
C>T
T>A
C>G
T>C
C>A
T>G
C
D
VAF
H3.1-K27M
n=18
H3.3-K27M
n=71
Subgroup frequency
logRatio
Subgroup
H3.1
H3.3
E
P < 0.01
P < 0.05
>3 (Co-occurence)
-log10(P-value)
> 3 (Mutually exclusive)

**Supplemental figure S7:** Summary of the mutations in DIPG – H3 status

**A-** Oncoplots depicting top 50 mutated genes sorted and ordered by decreasing frequency. The top barplot shows the total number of somatic non-synonymous coding mutations for each patient, while the right barplot indicates the frequency of mutations in each gene taking into consideration the union of Mutect and Varscan callings. Only somatic SNV with more than 5 supporting reads associated and a VAF exceeding 5% in both Mutect or Varscan calling are shown on the plot. Germinal alterations with more than 5 supporting reads associated and a VAF exceeding 40% are indicated as a small square. Two patients harbor a H3.3-K27M alteration with only 5 supporting reads and a sequencing depth of 44 and 49 for this gene and are indicated below as mutated for their H3 mutational status (asterisk). Tumors were grouped according to the cluster of CNV (A) or the histone H3-K27M status (C). The repartition of transversion and transition per patient is shown in the barplot below the heatmap.
**B-** Bar plot presenting for each gene, the percentage of cases presenting a somatic variation detected by Varscan on the left or Mutect on the right.
**D-** Violin plot of the mutated genes for which the VAF vary significantly between H3.1-K27M and H3.3-K27M tumors. Only genes presenting a difference in median VAF exceeding 10 % between the 2 subgroups and mutated in at least 2 patients in each subgroup (with a VAF >5%) are reported. The median VAF per gene is shown with the number of mutated patients in each condition indicated as the frequency are colored in dark green for H3.1-K27M and light green for H3.3-K27M.
**E-** Mutual exclusivity analysis. Pairwise Fishers exact tests are calculated for each of the 50 most commonly altered genes in DIPG. Log10-transformed pvalue are plotted in a brown to blue color scheme for co-occurence or mutual exclusivity respectively. * adjusted $p<0.01$, · adjusted p<0.05.

Figure S8

A

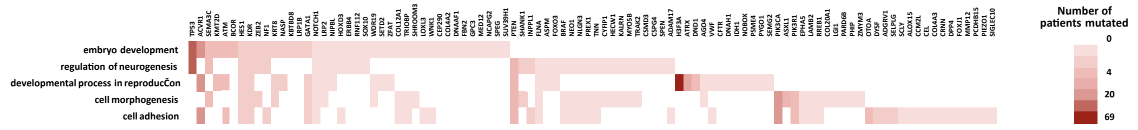

embryo development
regulation of neurogenesis
developmental process in reproduction
cell morphogenesis
cell adhesion
Number of patients mutated


B

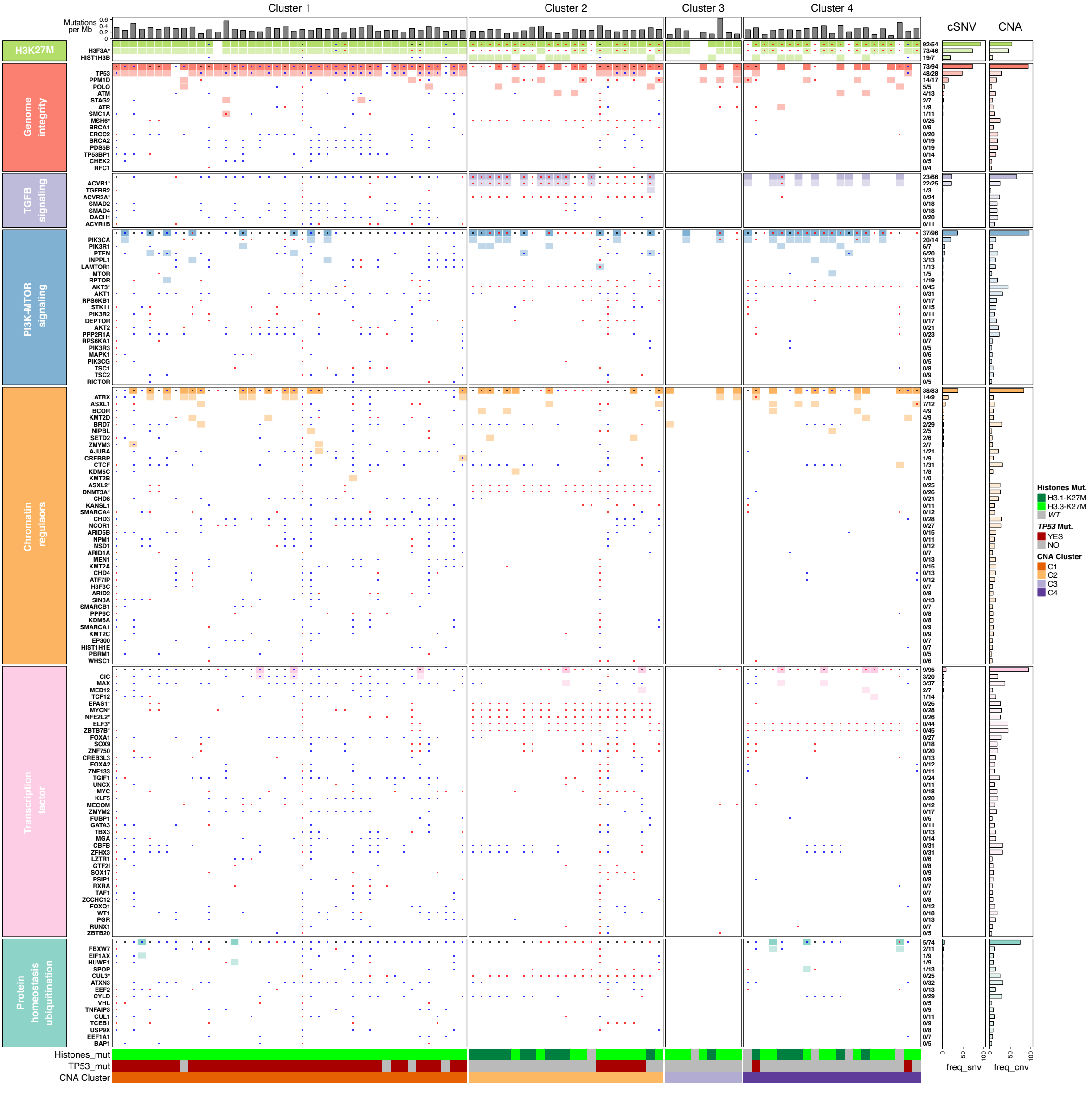

Cluster 1
Cluster 2
Cluster 3
Cluster 4
Mutations per Mb
cSNV
CNA
H3K27M
Genome integrity
TGFB signaling
PI3K-MTOR signaling
Chromatin regulaors
Transcription factor
Protein homeostasis ubiquitination
Histones_mut
TP53_mut
CNA Cluster
freq_snv
freq_cnv
Histones Mut.
H3.1-K27M
H3.3-K27M
WT
TP53 Mut.
YES
NO
CNA Cluster
C1
C2
C3
C4

**Supplemental figure S8:** Integrated pathway analysis of DIPG.

**A-** Biological process enrichment analysis of DIPG mutated genes.
The most significantly enriched (FDR adjusted Benjamini and Hodchberg from 1.79 E-04 to 8.47 E-5) and representative Gene Ontology categories from the list of genes mutated in at least two distinct patients of the entire cohort are presented. The heatmap reflect the number of patients presenting alteration.
**B-**Oncoprint representation of an integrated annotation of somatic mutations and DNA copy number changes affecting specific pathways. Point mutations are shown as a color rectangles, gains and losses are indicated as red and blue dots respectively. In case both gains and loss of a given gene are found in the cohort, the dot is indicated in black. Histone, *TP53* mutational status as well as their belonging to one molecular subgroup (I to IV) are indicated below the oncoprint. On the right the percentage of patients presenting SNV or copy-number alteration at gene-level or pathway-level are indicated and shown as a bar-plot. Genes located on chromosome 1q and chromosome 2 are indicated by an asterisk as they are frequently subjected to CNAs. Also, four patients present CNA in the majority of the genes depicted, resulting from a relative bad quality of DNA samples.
